# Deciphering Distinct Genetic Risk Factors for FTLD-TDP Pathological Subtypes via Whole-Genome Sequencing

**DOI:** 10.1101/2024.06.24.24309088

**Authors:** Cyril Pottier, Fahri Küçükali, Matt Baker, Anthony Batzler, Gregory D. Jenkins, Marka van Blitterswijk, Cristina T. Vicente, Wouter De Coster, Sarah Wynants, Pieter Van de Walle, Owen A. Ross, Melissa E. Murray, Júlia Faura, Stephen J. Haggarty, Jeroen GJ. van Rooij, Merel O. Mol, Ging-Yuek R. Hsiung, Caroline Graff, Linn Öijerstedt, Manuela Neumann, Yan Asmann, Shannon K. McDonnell, Saurabh Baheti, Keith A. Josephs, Jennifer L. Whitwell, Kevin F. Bieniek, Leah Forsberg, Hilary Heuer, Argentina Lario Lago, Ethan G. Geier, Jennifer S. Yokoyama, Alexis P. Oddi, Margaret Flanagan, Qinwen Mao, John R. Hodges, John B. Kwok, Kimiko Domoto-Reilly, Matthis Synofzik, Carlo Wilke, Chiadi Onyike, Bradford C. Dickerson, Bret M. Evers, Brittany N. Dugger, David G. Munoz, Julia Keith, Lorne Zinman, Ekaterina Rogaeva, EunRan Suh, Tamar Gefen, Changiz Geula, Sandra Weintraub, Janine Diehl-Schmid, Martin R. Farlow, Dieter Edbauer, Bryan K. Woodruff, Richard J. Caselli, Laura L. Donker Kaat, Edward D. Huey, Eric M. Reiman, Simon Mead, Andrew King, Sigrun Roeber, Alissa L. Nana, Nilufer Ertekin-Taner, David S. Knopman, Ronald C. Petersen, Leonard Petrucelli, Ryan J. Uitti, Zbigniew K. Wszolek, Eliana Marisa Ramos, Lea T. Grinberg, Maria Luisa Gorno Tempini, Howard J. Rosen, Salvatore Spina, Olivier Piguet, Murray Grossman, John Q. Trojanowski, Dirk C. Keene, Jin Lee-Way, Johannes Prudlo, Daniel H. Geschwind, Robert A. Rissman, Carlos Cruchaga, Bernardino Ghetti, Glenda M. Halliday, Thomas G. Beach, Geidy E. Serrano, Thomas Arzberger, Jochen Herms, Adam L. Boxer, Lawrence S. Honig, Jean P. Vonsattel, Oscar L. Lopez, Julia Kofler, Charles L. White, Marla Gearing, Jonathan Glass, Jonathan D. Rohrer, David J. Irwin, Edward B. Lee, Vivianna Van Deerlin, Rudolph Castellani, Marsel M. Mesulam, Maria C. Tartaglia, Elizabeth C. Finger, Claire Troakes, Safa Al-Sarraj, Bruce L. Miller, Harro Seelaar, Neill R. Graff-Radford, Bradley F. Boeve, Ian RA. Mackenzie, John C. van Swieten, William W. Seeley, Kristel Sleegers, Dennis W. Dickson, Joanna M. Biernacka, Rosa Rademakers

## Abstract

Frontotemporal lobar degeneration with neuronal inclusions of the TAR DNA-binding protein 43 (FTLD-TDP) is a fatal neurodegenerative disorder with only a limited number of risk loci identified. We report our comprehensive genome-wide association study as part of the International FTLD-TDP Whole-Genome Sequencing Consortium, including 985 cases and 3,153 controls, and meta-analysis with the Dementia-seq cohort, compiled from 26 institutions/brain banks in the United States, Europe and Australia. We confirm *UNC13A* as the strongest overall FTLD-TDP risk factor and identify *TNIP1* as a novel FTLD-TDP risk factor. In subgroup analyses, we further identify for the first time genome-wide significant loci specific to each of the three main FTLD-TDP pathological subtypes (A, B and C), as well as enrichment of risk loci in distinct tissues, brain regions, and neuronal subtypes, suggesting distinct disease aetiologies in each of the subtypes. Rare variant analysis confirmed *TBK1* and identified *VIPR1*, *RBPJL*, and *L3MBTL1* as novel subtype specific FTLD-TDP risk genes, further highlighting the role of innate and adaptive immunity and notch signalling pathway in FTLD-TDP, with potential diagnostic and novel therapeutic implications.

## Introduction

Frontotemporal lobar degeneration (FTLD) is one of the leading causes of dementia in individuals younger than 65 years but can also affect individuals later in life. The predominant clinical presentations of FTLD are behavior and language dysfunction resulting in behavioral variant frontotemporal dementia (bvFTD)^1^, semantic variant primary progressive aphasia (svPPA), or nonfluent variant primary progressive aphasia (nfvPPA)^2^. The diagnosis of FTLD can be established with certainty only with neuropathologic postmortem examination and is characterized neuropathologically with significant atrophy of the frontal and temporal lobes and accumulation of abnormal neuronal and/or glial inclusions upon immunohistochemical analysis. FTLD-TDP, characterized by neuronal and cytoplasmic aggregates of the DNA and RNA-binding protein TDP-43, is one of the two main pathological subtypes (the other being FTLD-Tau) and can be further classified into five FTLD-TDP subtypes (FTLD-TDP A-E) based on the distribution of the neuronal cytoplasmic TDP-43-positive inclusions and dystrophic neurites in the cortical layers^3,4^. In general, an accurate prediction of the underlying neuropathological FTLD subtype of individual patients constitutes a diagnostic challenge; however, a few clinicopathological correlations exist. Specifically, over 80% of patients clinically presenting with svPPA are diagnosed as FTLD-TDP C at autopsy^5^, and patients with bvFTD with concomitant amyotrophic lateral sclerosis (ALS) almost invariably present as FTLD-TDP B at autopsy^2,4,6^.

A small number of autosomal dominant genes and risk factors associated with FTLD-TDP have been reported^7–13^. The first FTLD-TDP genome-wide association study (GWAS) identified the *TMEM106B* locus (rs1990622), supporting lysosomal dysfunction in FTLD-TDP; however, this signal was strongly driven by *GRN* mutation carriers included in that study. Three additional FTLD-TDP loci, *UNC13A*, *DPP6* and *HLA*-*DQA2,* were identified in phase I of the International FTLD-TDP whole-genome sequencing (WGS) consortium and require replication in larger datasets^11^. Importantly, most FTLD-TDP patients are not yet genetically explained, and the relatively small sample size precluded rare variant analyses in phase I.

To replicate and identify new genetic risk factors, we doubled the original sample size of the FTLD-TDP WGS consortium by not only sequencing more pathologically confirmed FTLD-TDP cases but also including clinically defined FTLD subtypes enriched for specific FTLD-TDP pathological subtypes at autopsy. GWAS analyses of both common and rare variants, followed by comprehensive gene-prioritization, enrichment analyses, and co-localization studies identified novel FTLD-TDP risk loci, including novel risk genes and loci specific to FTLD-TDP pathological subtypes. Our study highlights similarities and differences between FTLD-TDP and other neurodegenerative diseases while unique biological processes in specific tissues, brain regions, and cell types were found to characterize individual FTLD-TDP pathological subtypes.

## Results

### GWAS analysis

#### Common variant genome-wide association study

To identify novel common FTLD-TDP genetic risk factors, we performed single variant GWAS using an additive disease risk model for 6,568,099 common variants in 985 patients and 3,153 controls free of neurodegenerative disorder that passed quality control (QC). Combining all patients (FTLD-TDP All) we identified one genome-wide significant signal at the *UNC13A* locus (rs8111424, OR=1.37, *P*=1.17×10^-8^). We also performed separate GWAS within the FTLD-TDP A, FTLD-TDP B, and FTLD-TDP C pathological subtypes (**Fig. 1**, **Tables 1-2**). The most significant locus identified in FTLD-TDP A was *GRN* (rs5848; OR=1.89, *P*=5.57×10^-9^). In phase I, this locus only reached genome-wide significance under an exploratory recessive model^11^, and also now, the recessive model provided an even stronger association (OR=4.12, *P*=8.28×10^-15^). We further detected 3 additional new genome-wide significant loci in FTLD-TDP A: *TINAG* (rs138698596), *MZT1* (rs138959102) and *FARP2* (rs886815). In FTLD-TDP B, we detected a genome-wide significant association at the *UNC13A* locus (rs12973192). The lead variant rs12973192 is in linkage disequilibrium (LD) with rs8111424 (D’=1; r^2^=0.43) identified in the FTLD-TDP All analysis. We further detected 3 new genome-wide significant risk loci in FTLD-TDP B: *TNIP1* (rs871269), *RCL1* (rs7674221), *PDS5B* (rs5277499), and one in FTLD-TDP C *C19orf52* (also known as *TIMM29*, rs576561313).

**Figure 1:**
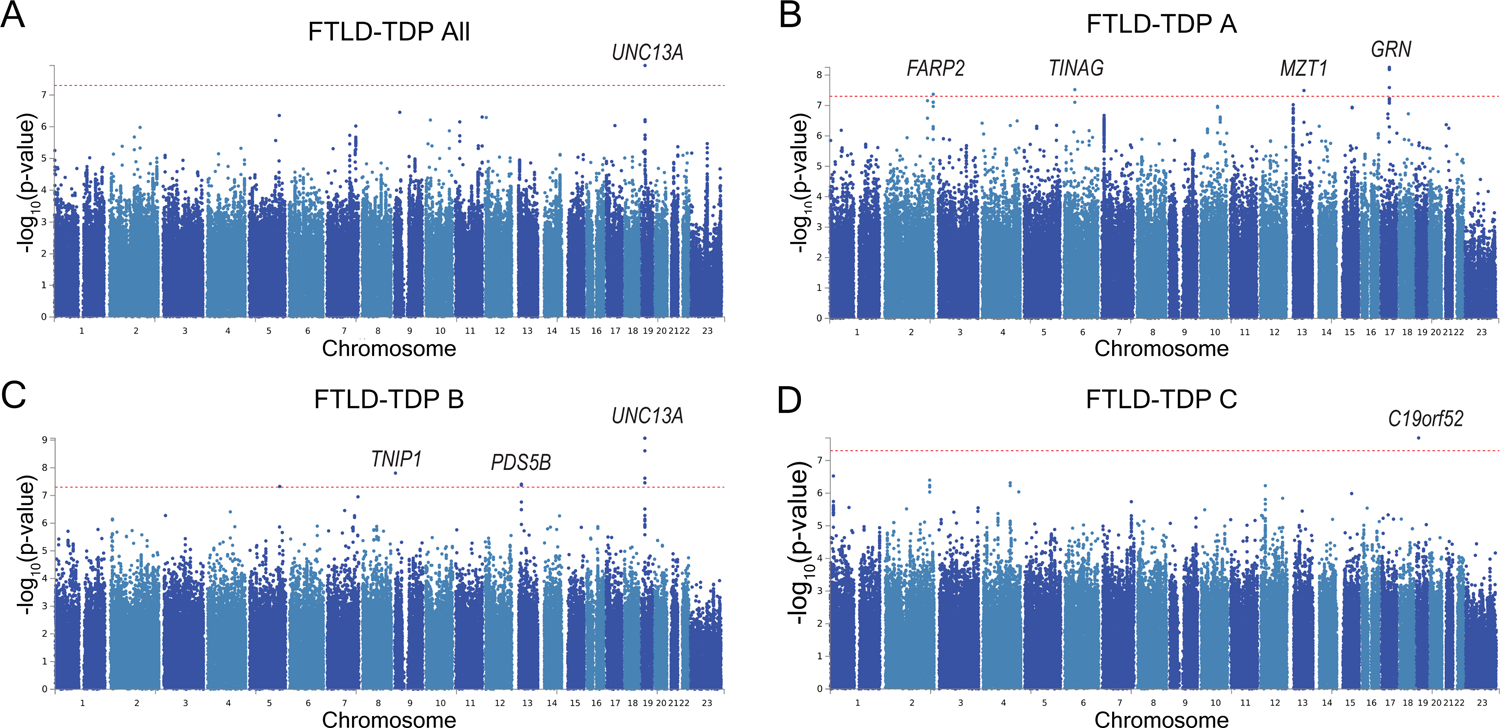
Genome-wide association study on common variants. **A** Manhattan plot of the FTLD-TDP All patients versus controls association study. **B** Manhattan plot of the FTLD-TDP A patients versus controls association study. **C** Manhattan plot of the FTLD-TDP B patients versus controls association study. **D** Manhattan plot of the FTLD-TDP C patients versus controls association study. The red-dotted line represents the genome-wide significance level (p = 5×10^-8^).

**Table 1:**
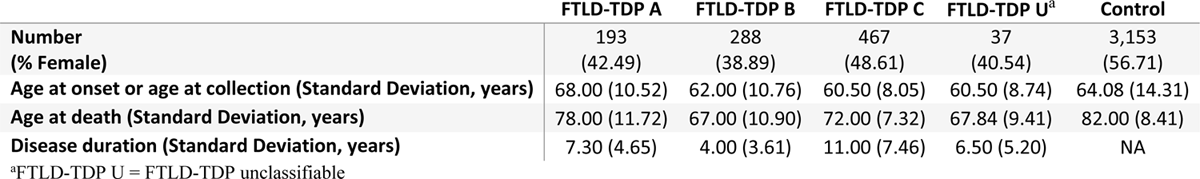
Demographics after quality control.

|  | FTLD-TDP A | FTLD-TDP B | FTLD-TDP C | FTLD-TDP U<sup>a</sup> | Control |
| --- | --- | --- | --- | --- | --- |
| **Number** | 193 | 288 | 467 | 37 | 3,153 |
| **(% Female)** | (42.49) | (38.89) | (48.61) | (40.54) | (56.71) |
| **Age at onset or age at collection (Standard Deviation, years)** | 68.00 (10.52) | 62.00 (10.76) | 60.50 (8.05) | 60.50 (8.74) | 64.08 (14.31) |
| **Age at death (Standard Deviation, years)** | 78.00 (11.72) | 67.00 (10.90) | 72.00 (7.32) | 67.84 (9.41) | 82.00 (8.41) |
| **Disease duration (Standard Deviation, years)** | 7.30 (4.65) | 4.00 (3.61) | 11.00 (7.46) | 6.50 (5.20) | NA |

<sup>a</sup>FTLD-TDP U = FTLD-TDP unclassifiable

**Table 2:**
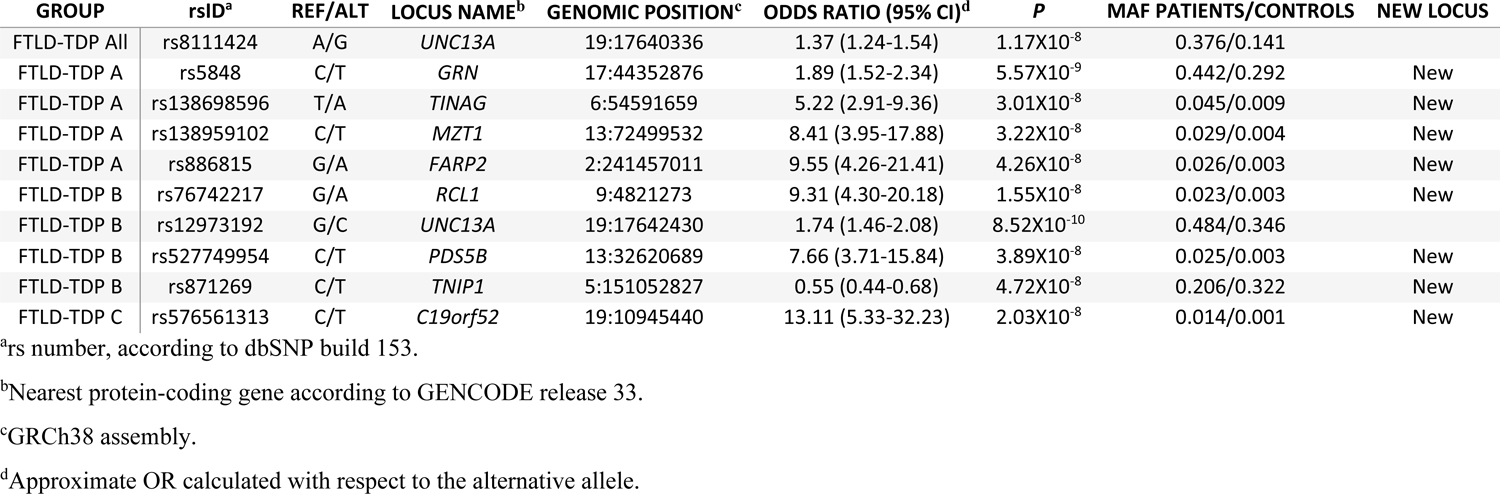
Top variants associated with disease status.

| GROUP | rsID<sup>a</sup> | REF/ALT | LOCUS NAME<sup>b</sup> | GENOMIC POSITION<sup>c</sup> | ODDS RATIO (95% CI)<sup>d</sup> | P | MAF PATIENTS/CONTROLS | NEW LOCUS |
| --- | --- | --- | --- | --- | --- | --- | --- | --- |
| FTLD-TDP All | rs8111424 | A/G | *UNC13A* | 19:17640336 | 1.37 (1.24-1.54) | 1.17X10<sup>-8</sup> | 0.376/0.141 |  |
| FTLD-TDP A | rs5848 | C/T | *GRN* | 17:44352876 | 1.89 (1.52-2.34) | 5.57X10<sup>-9</sup> | 0.442/0.292 | New |
| FTLD-TDP A | rs138698596 | T/A | *TINAG* | 6:54591659 | 5.22 (2.91-9.36) | 3.01X10<sup>-8</sup> | 0.045/0.009 | New |
| FTLD-TDP A | rs138959102 | C/T | *MZT1* | 13:72499532 | 8.41 (3.95-17.88) | 3.22X10<sup>-8</sup> | 0.029/0.004 | New |
| FTLD-TDP A | rs886815 | G/A | *FARP2* | 2:241457011 | 9.55 (4.26-21.41) | 4.26X10<sup>-8</sup> | 0.026/0.003 | New |
| FTLD-TDP B | rs76742217 | G/A | *RCL1* | 9:4821273 | 9.31 (4.30-20.18) | 1.55X10<sup>-8</sup> | 0.023/0.003 | New |
| FTLD-TDP B | rs12973192 | G/C | *UNC13A* | 19:17642430 | 1.74 (1.46-2.08) | 8.52X10<sup>-10</sup> | 0.484/0.346 |  |
| FTLD-TDP B | rs527749954 | C/T | *PDS5B* | 13:32620689 | 7.66 (3.71-15.84) | 3.89X10<sup>-8</sup> | 0.025/0.003 | New |
| FTLD-TDP B | rs871269 | C/T | *TNIP1* | 5:151052827 | 0.55 (0.44-0.68) | 4.72X10<sup>-8</sup> | 0.206/0.322 | New |
| FTLD-TDP C | rs576561313 | C/T | *C19orf52* | 19:10945440 | 13.11 (5.33-32.23) | 2.03X10<sup>-8</sup> | 0.014/0.001 | New |

<sup>a</sup>rs number, according to dbSNP build 153.
<sup>b</sup>Nearest protein-coding gene according to GENCODE release 33.
<sup>c</sup>GRCh38 assembly.
<sup>d</sup>Approximate OR calculated with respect to the alternative allele.

In order to prioritize risk genes and identify possible biological mechanisms, we applied a range of variant annotation and molecular quantitative trait loci (QTL)-GWAS integration analyses as previously described^14^ (**Supplementary Tables 2-13**). We integrated different levels of evidence using a weighting scheme and obtained a weighted sum of the hits in different subcategories for each gene. We grouped candidate risk genes in genome-wide significant loci and in subthreshold loci and prioritized them at two levels of confidence for being a likely risk gene as tier 1 (higher confidence) and tier 2 (lower confidence).

The gene prioritization analyses nominated a total of 59 tier 1 and 246 tier 2 genes in 258 different loci for 4 different GWAS analyses (**Fig. 2, Supplementary Table 3, Supplementary** Fig. 2-5). Our results showed that the nearest protein-coding genes were prioritized as tier 1 (*n*=8) and tier 2 (*n*=1) risk genes in the genome-wide significant loci for the distinct FTLD-TDP subtypes. Of the 8 tier 1 prioritized genes, 3 were found in common variant loci where molecular QTL-GWAS analyses aided their prioritization (**Fig. 2**). First, in locus A4, *GRN* was prioritized through consistent expression QTL (eQTL) domain hits in bulk brain regions (eQTL *P*_ROSMAP DLPFC_=6.32×10^-38^ and beta_ROSMAP DLPFC_=-0.25, eQTL colocalization (coloc) PPH4s of 81.8%-99.7%, and fine-mapped [posterior inclusion probability being 100%] expression transcriptome wide association study [eTWAS] associations with *P* from 1.74×10^-8^ to 5.15×10^-9^ and Z-scores from −5.63 to −5.84) and in oligodendrocytes (cell type specific eQTL (ct-eQTL) coloc PPH4=90%) where genetic downregulation of *GRN* gene expression was associated with the FTLD-TDP A risk signal (**Supplementary Tables 5, 6**, and **11**), which was also observed in brain proteome-wide association study (PWAS) with the same effect direction for the FTLD-TDP A risk (*P*_ROSMAP DLPFC_ = 3.32×10^-6^, Z_ROSMAP DLPFC_ = −4.65, **Supplementary Table 13**). Second, in locus B1, *TNIP1* was prioritized because the minor allele was associated with decreased *TNIP1* expression (*P*_ROSMAP DLPFC_ = 2.40×10^-4^, beta_ROSMAP DLPFC_ = −0.10), the GWAS signal colocalized with a microglia splicing QTL (sQTL) associated with *TNIP1* chr5:151032383-151035002 known splice junction (coloc PPH4=82.2%) and because the methylation QTL (mQTL) variants for cg03340667, a CpG ∼3.7 kb upstream of the transcription start site (TSS) of the canonical transcript of *TNIP1*, colocalized with the GWAS variants (coloc PPH4=70%) in dorsolateral prefrontal cortex (DLPFC) (**Supplementary Tables 5, 7**, and **8**). Third, in locus B4, *UNC13A* was prioritized through an eQTL-GWAS colocalization in temporal cortex (coloc PPH4=81.82%, **Supplementary Table 6**). Furthermore, beyond genome-wide significant loci, we identified additional candidate prioritized risk genes in subthreshold regions through molecular QTL-GWAS coloc and TWAS analyses, one important example being *TMEM106B* as the prioritized risk gene in locus A_S14. The FTLD-TDP A GWAS signal near *TMEM106B* colocalized with eQTL variants regulating *TMEM106B* gene expression in bulk brain regions (eQTL coloc PPH4s=81.40% in MayoRNASeq temporal cortex and 89.66% in GTEx brain cortex, **Supplementary Table 6**). We also observed a significant eTWAS association in GTEx cortex (*P*=4.13×10^-7^, Z=-5.06), together with a significant PWAS hit (*P*=2.01×10^-8^, Zscore=5.61) **(Supplementary Tables 11, 13**). Finally, a significant hit in splicing TWAS in cortex (sTWAS, *P*=6.66×10^-7^, Z=-4.97) predicted a decreased preference for the *TMEM106B* splice junction chr7:12224385-12229679 with the increased FTLD-TDP A GWAS risk, while we also observed methylation QTL (mQTL) coloc hits for two CpGs for *TMEM106B* (∼500 bp upstream cg23422036 coloc PPH4=94.25% and intronic cg09613507 coloc PPH4=94.09%) (**Supplementary Tables 12, 8**). We summarized our gene prioritization results in **Fig. 2** for the genome-wide significant loci and a selection of the suggestive loci (with genes having GWAS evidence of *P* ≤ 5×10^-6^ in 1 Mb extended regions), and full results are presented in **Supplementary Tables 3-13 and Supplementary** Fig. 2-5.

**Figure 2:**
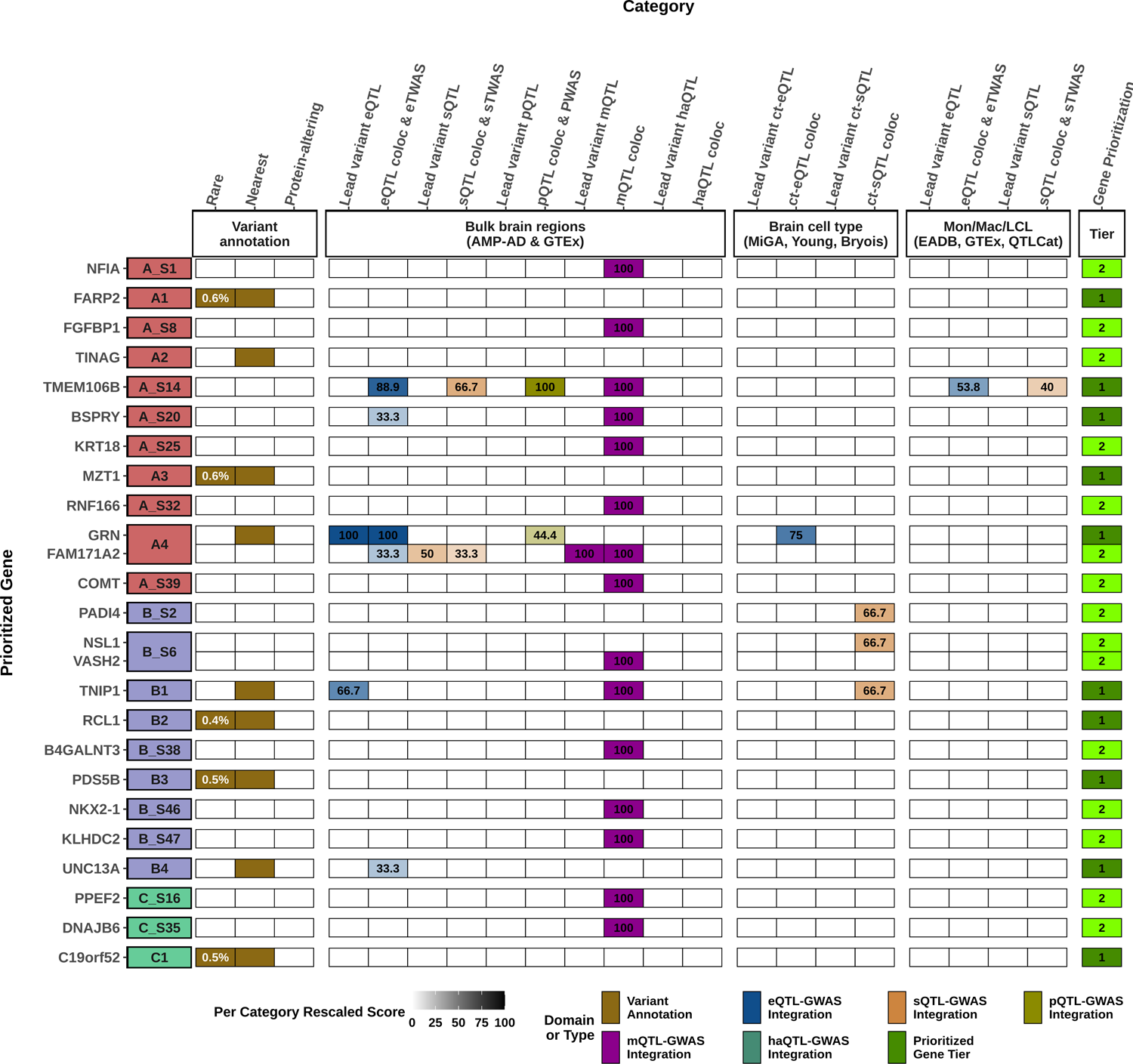
Gene prioritization results for FTLD-TDP subgroups. A visual summary of weighted evidence category scores for the prioritized genes within genome-wide significant and subthreshold loci with candidate genes whose 1 Mb extended gene coordinates contain a minimal GWAS *P* evidence of ≤5×10^-6^ in related FTLD subtype-specific GWAS summary statistics. Using the gene prioritization strategy in these selected loci, we prioritized a total of 25 genes in 23 loci at two different confidence levels (10 tier 1 and 15 tier 2 prioritized genes). The leftmost squares which are colored in red for FTLD-TDP A, in blue for FTLD-TDP B, and in green for FTLD-TDP C specific analyses indicate the locus index numbers which contain additional “_S” patterns for the subthreshold loci, whereas others indicate the genome-wide significant loci. The types of evidence for each category are colored according to the six different domains to which they belong. Weighted scores for each evidence category are rescaled to a 0–100 scale based on the maximum score a candidate gene can obtain from a category (see **Supplementary Table 2**). The darker colors represent higher scores in categories, while tier 1 prioritized genes are displayed in dark green and tier 2 prioritized genes are displayed in light green. Only tier 1 and tier 2 genes are shown for each locus, and all candidate genes considered and scored can be found in **Supplementary Table 3**. MAFs (based on gnomAD v4 non-Finnish European samples) and CADD (v1.7) PHRED scores for rare and/or protein-altering rare variants are labeled in white within the respective squares. eQTL, expression QTL; sQTL, splicing QTL; mQTL, methylation QTL; pQTL, protein-expression QTL; haQTL, histone acetylation QTL; coloc, colocalization; eTWAS, expression transcriptome-wide association study; sTWAS, splicing transcriptome-wide association study; PWAS, proteome-wide association study; Mon. Mac., monocytes and macrophages; LCL, lymphoblastoid cell line; QTLCat, The eQTL Catalogue.

Next, we performed gene ontology analyses on tier1 prioritized genes. The most significant term in the nominated genes in FTLD-TDP All was positive regulation of defense response to bacterium (*P*=3.21×10^-5^). Lysosomal function appeared to be strongly affected in FTLD-TDP A with several genes such as *GRN* and *TMEM106B* (lysosomal organization GO term, *P*=4.12×10^-4^) as well as cathepsin B (*CTSB*). We further detected enriched terms for retrograde transport in FTLD-TDP B (*P*=2.21×10^-3^) driven by *DENND2A* and *VPS53* genes and for excitatory postsynaptic potential in FTLD-TDP C (p=1.48×10^-3^) driven by *DMPK* and *P2RX5* genes (**Fig. 3, Supplementary Table 14**). Importantly, except for lysosomal transport, no terms overlapped between subtypes of FTLD-TDP, suggesting mostly distinct genetic etiologies in the different FTLD-TDP groups.

**Figure 3:**
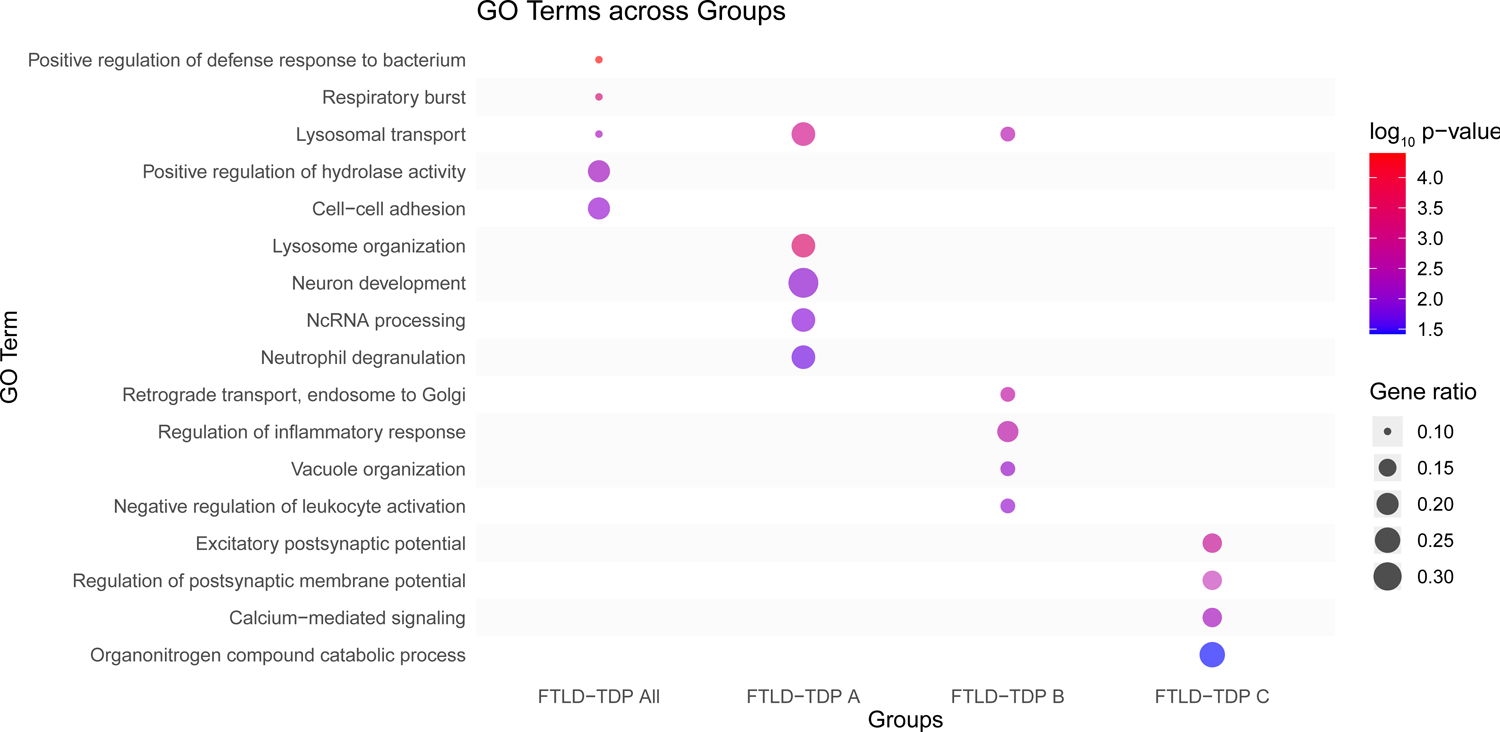
Top 5 Gene Ontology terms enriched in FTLD-TDP subgroups. Hierarchical GO analysis of biological process terms considering genes in genetic loci prioritized for FTLD-TDP All, FTLD-TDP A, FTLD-TDP B and FTD-TDP C.

To further characterize genetic factors associated with FTLD-TDP, we performed gene-based analyses on common variants with *P*<10^-5^ using MAGMA. Analyses of FTLD-TDP All did not yield exome-wide significant loci; however, FTLD-TDP A showed exome-wide significant signals for the two genes located at the *GRN* locus (*FAM171A2*, *ITGA2B*) and for *TMEM106B* (*P*=4.74×10^-7^). The *TMEM106B* signal was driven by the rs10281425 variant (OR=0.54, *P*=2.12×10^-7^). No exome-wide significant signal was detected for the other two FTLD-TDP pathological subtypes.

#### Expression of the top suggestive signals in cell types and brain regions

To find tissues and cell types for which gene expression profiles were enriched for genes within FTLD-risk loci, we combined gene-based association statistics calculated using MAGMA with gene expression patterns from the Genotype–Tissue Expression (GTEx) project in a gene set enrichment analysis. We observed an enrichment in genes expressed in brain tissue (cerebellum, frontal cortex, and cortex) in FTLD-TDP A and B. This was strikingly different from the signature observed in FTLD-TDP C for which significant enrichment was only detected in non-central nervous system tissue, in particular small intestine terminal ileum (**Fig. 4, Supplementary Table 15**). We also compared FTLD gene expression loci with similar data obtained from Alzheimer’s disease and related disorders (ADRD)^14^ and ALS GWAS^15^. FTLD-TDP subtypes presented with a distinct genetic signature as compared to these related disorders highlighting the importance of regional specificity in FTLD-TDP.

**Figure 4:**
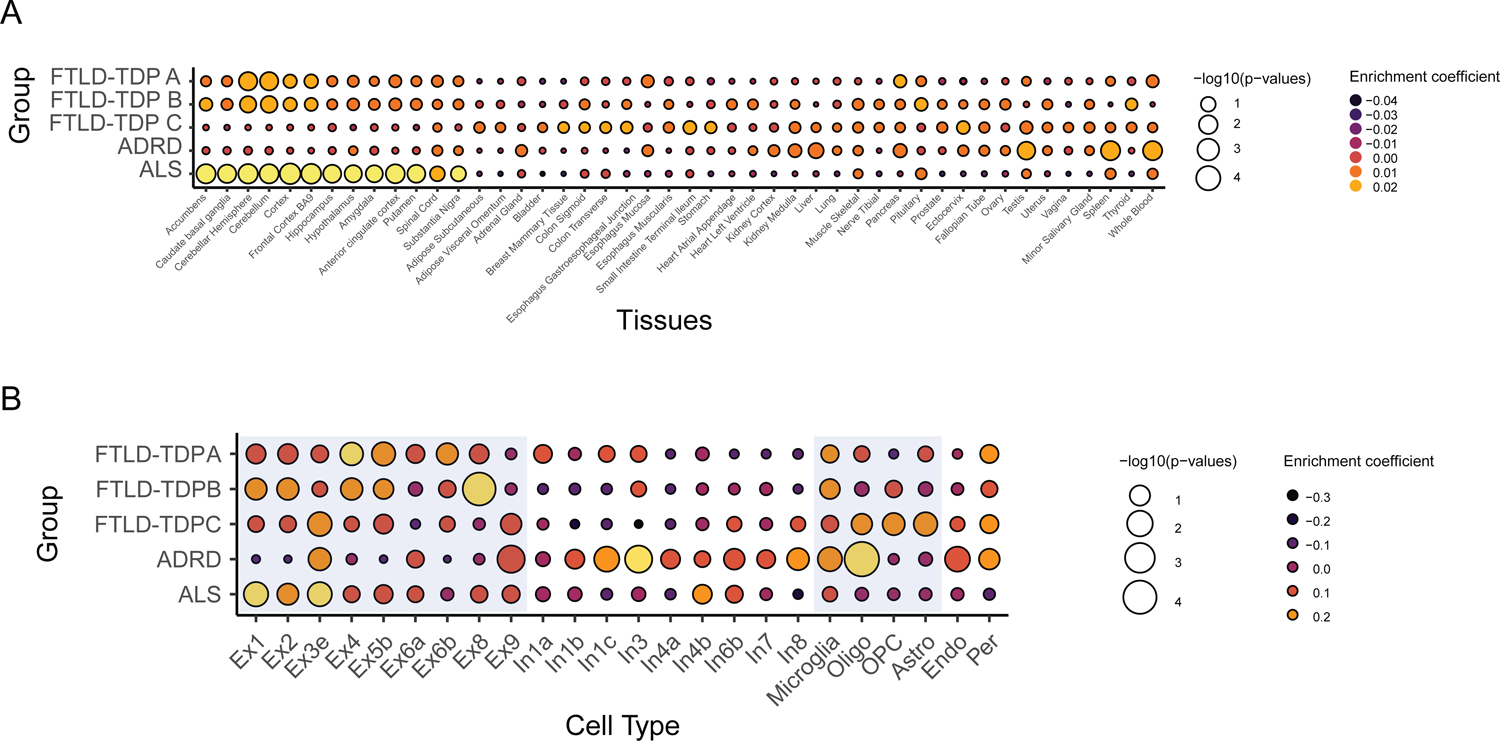
Enrichment of brain regions and cell types in FTLD subgroups. **A** Enrichment of genes in multiple tissues, including 13 brain regions, and based on GTEX data in FTLD subgroups, ADRD and ALS. Color represents the enrichment coefficient, and size indicates two-sided −log10 (FDR adjusted *P*s) of enrichment obtained by the linear regression model in the MAGMA gene property analysis. **B** Central nervous system cell type enrichment analyses in FTLD subgroups, ADRD and ALS. Color represents the enrichment coefficient, and size indicates two-sided −log10 (FDR adjusted *P*s) of enrichment obtained by the linear regression model in the MAGMA gene property analysis. Excitatory neurons and glial cells are highlighted in blue. Excitatory and inhibitory neurons from the PsychENCODE dataset were labeled based on their transcriptional profile from 1 to 8. Asterisks denote brain regions or cell types enriched with FDR *P*<0.05. Cx, cortex; Ex*, Excitatory neuron, In*, inhibitory neurons; Oligo, oligodendrocytes; OPCs, oligodendrocyte progenitor cells; Astro, astrocytes; Endo, endothelial cells; Per, pericytes.

We subsequently queried PsychENCODE frontal-cortex single-cell RNA-seq datasets of human-derived brain samples to specify further which brain-specific enriched cell types express the genetic loci associated with FTLD-TDP risk (**Fig. 4, Supplementary Table 16**). We observed a significant enrichment in genes expressed in excitatory neurons for FTLD-TDP A loci (Ex4 *P*=3.55×10^-2^, Ex5b *P*=2.72×10^-2^), and FTLD-TDP B loci (Ex8 *P*=1.27×10^-4^), while no other cell type reached significance. While FTLD-TDP C loci were also significantly enriched in genes expressed in excitatory neurons (Ex3e *P*=2.10×10^-2^), they were additionally enriched in genes expressed in astrocytes and oligodendrocyte progenitor cells (*P*=4.69×10^-2^, *P*=2.53×10^-2^). Genes expressed in microglia were enriched only in ADRD gene loci (*P*=1.90×10^-2^). Overall, loci comprising genes expressed in excitatory neurons were enriched in the three FTLD-TDP subtypes with stronger specificity for specific neuron types in each FTLD-TDP subtype as compared to what was observed for ALS gene loci.

#### UNC13A and TNIP1 loci

To provide further support for the identified FTLD-TDP risk loci, we performed a meta-analysis of our FTLD-TDP cohort with the Dementia-seq study (phs001963.v1.p1) that includes 2,102 clinical FTLD patients and 1,748 controls. Given that this cohort lacks details on the FTLD pathology underlying each patient, pathological subgroup analyses could not be performed. Meta-analysis confirmed *UNC13A* and identified the new *TNIP1* locus as genome-wide significantly associated with FTLD (*P*_rs12973192_=8.85×10^-10^; *P*_rs871269_=3.42×10^-8^, respectively). Note that the most significant single nucleotide variant (SNV) at the *UNC13A* locus was rs12608932 (p=9.13×10^-11^), in strong LD with rs12973192 (r^2^=0.96 D’=0.99).

Both *UNC13A* and *TNIP1* were previously associated with other neurodegenerative diseases^14,16^. Colocalization analyses showed that our *UNC13A* signal was shared with ALS (coloc PPH4=95.71%), strongly confirming the genetic overlap between both diseases (**Fig. 5A**). Interestingly, for *TNIP1* we found strong colocalization with the ADRD association signal^14,16^ (coloc PPH4=99.2%) while its colocalization with ALS was weaker (71.5%), which was confirmed in a specificity analysis (coloc PPH4=20.1%, for p12=10e-06), possibly reflecting multiple independent association signals in FTLD in this locus (**Fig. 5B-C**).

**Figure 5:**
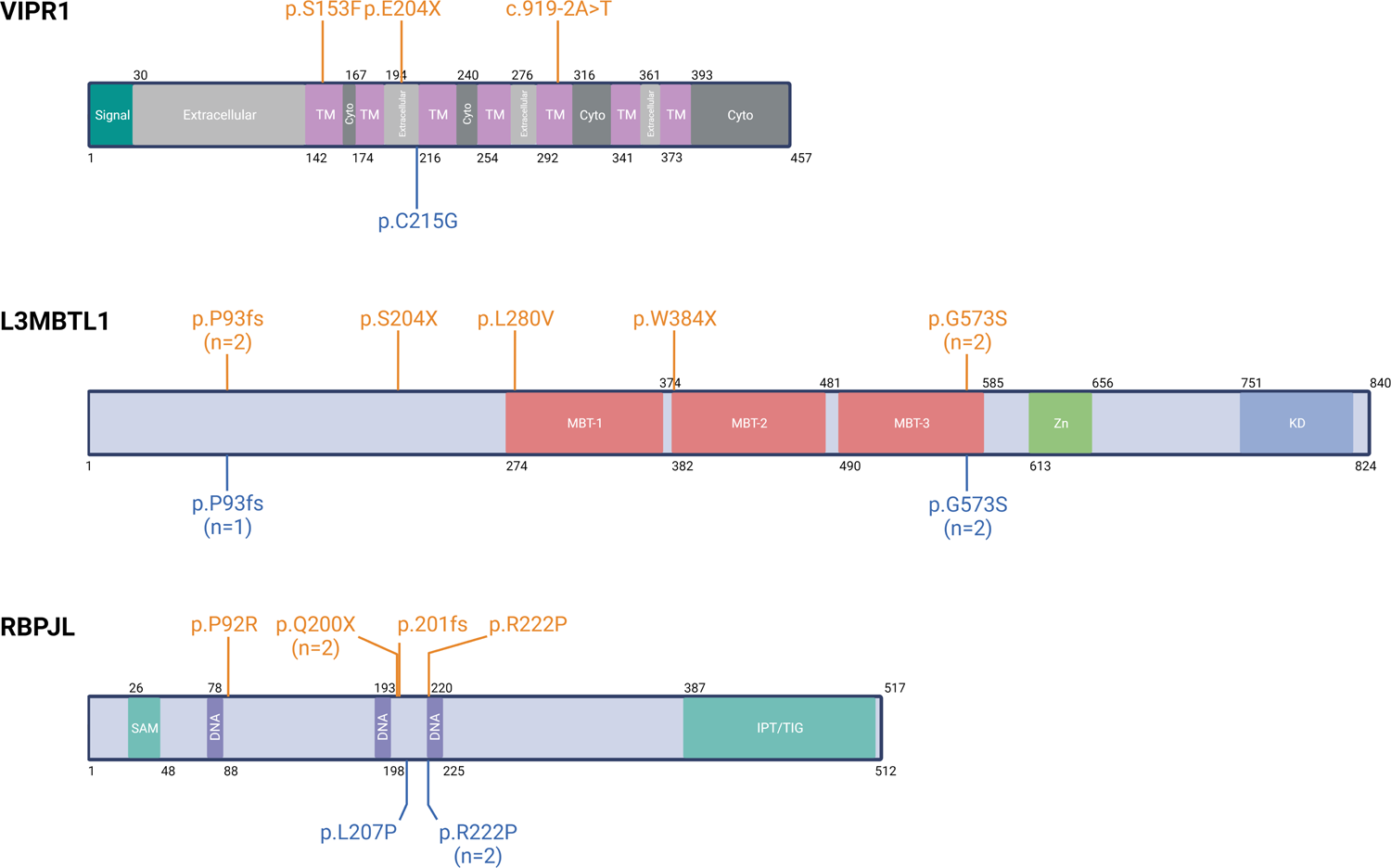
Locus zoom plots for *UNC13A* and *TNIP1* loci. **A** Genetic colocalization between the *UNC13A* locus in FTLD-TDP (meta-analysis) and ALS signal. **B** Genetic colocalization between the *TNIP1* locus in FTLD-TDP (meta-analysis) and ALS. **C** Genetic colocalization between the *TNIP1* locus in FTLD-TDP (meta-analysis) and ADRD. For **A**, **B** and **C**, chromosome position is located on the x axis and −log10(*P*) is represented on the y axis. Each dot represents a SNV tested in the dataset for its association with disease status. Purple diamonds are the index SNVs reported. Linkage disequilibrium with index SNV is indicated by r^2^.

#### Rare variant analysis

To identify genes carrying rare variants contributing to FTLD-TDP, we performed a burden test in genes with variants likely to affect protein function. In the overall FTLD-TDP cohort, no exome wide significant gene was detected (**Supplementary Table 17**). We did detect five exome-wide significant signals within FTLD-TDP pathological subtypes (**Table 3, Supplementary Table 18**). *TBK1* was associated with disease status in FTLD-TDP A and B (*P*=1.27×10^-11^, *P*=3.17×10^-12^, respectively). The signal was driven by 3 carriers in FTLD-TDP A patients (3/193=1.5%) and 5 carriers in FTLD-TDP B patients (5/288=1.7%). We also detected an enrichment in rare variants in *VIPR1* in FTLD-TDP B (*P*=4.65e-07, 3/288 FTLD-TDP B and 1/3153 control; **Fig. 6**) and 2 exome wide significant signals in FTLD-TDP C *L3MBTL1* (*P*=2.87×10^-7^, 8/467 FTLD-TDP C and 3/3153 controls) and *RBPJL* (*P*=6.39×10^-7^, 5/467 FTLD-TDP C and 3/3153 controls). Weighted gene coexpression network analysis using the ROSMAP dataset and the BrainExp database^17^ revealed that *L3MBTL1* and *RBPJL* belonged to the same module (yellow, *P_L3MBTL1_*=1.32×10^-45^, *P_RBPJL_*=1.00×10^-79^; **Supplementary** Figure 6) that is enriched in neuroactive ligand-receptor interaction and the cytokine-cytokine receptor interaction gene-ontology terms (*P*_FDR_=3.7×10^-12^, *P*_FDR_=5.8×10^-12^, respectively). While expression of *L3MBTL1* was throughout the central nervous system cells, *RBPJL* expression was restricted to inhibitory neurons and in particular to Parvalbumin neurons (**Supplementary** Figure 6).

**Figure 6:**
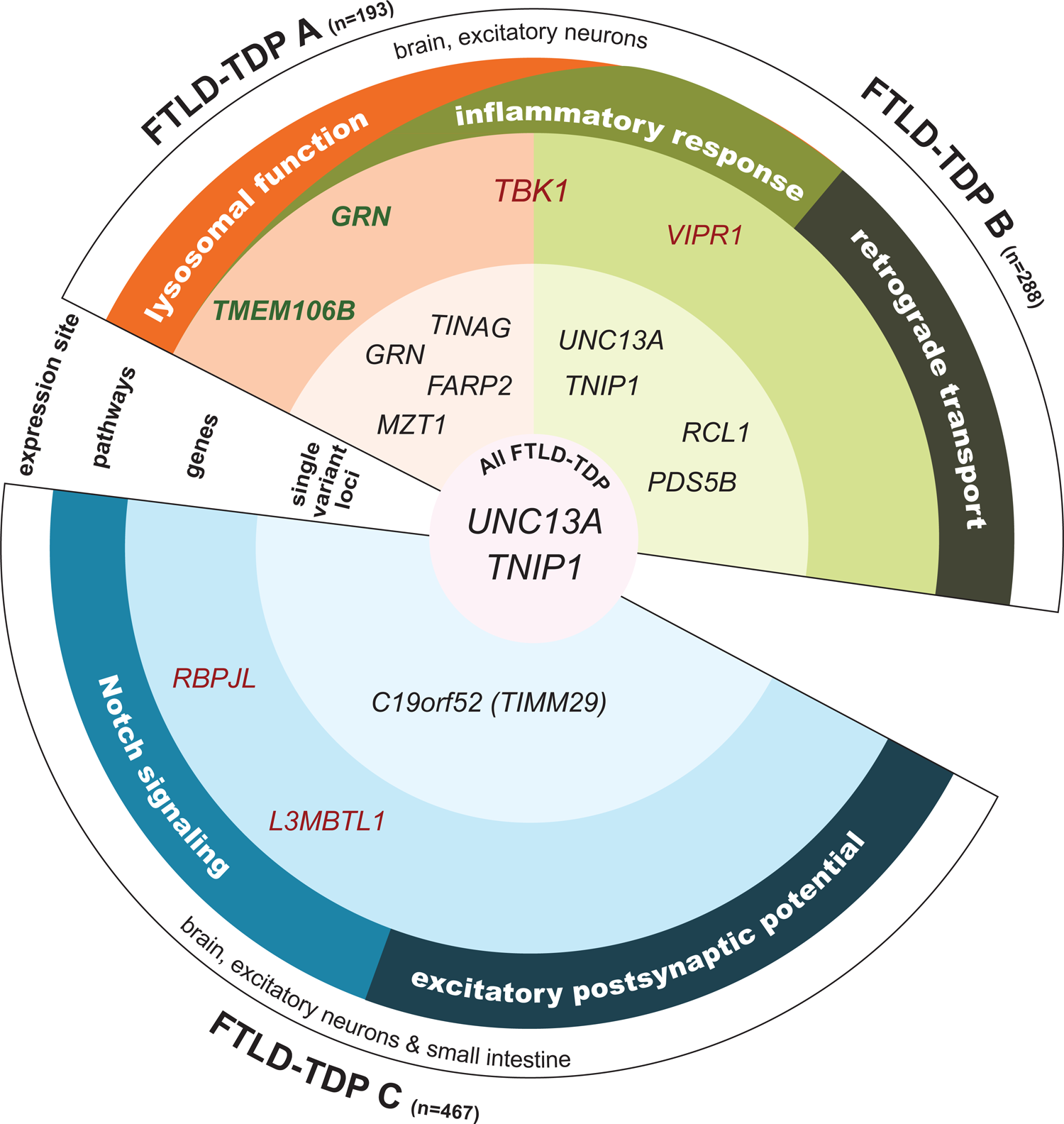
Rare loss of function and predicted pathogenic variants in proteins associated with FTLD. Schematic representation of VIPR1, L3MBTL1 and RBPL protein structure (source Uniprot) showing a map of nonsense, splicing, frameshift and missense with a REVEL score>0.75 rare variants in patients and controls. Variants identified in patients are colored in orange, variants identified in controls are colored in blue. n=number of carriers.

**Table 3:** Genes harboring rare variants associated with FTLD-TDP.

| GROUP | GENE NAME | NUMBER OF PATIENTS | MAF<sup>a</sup> IN PATIENTS | NUMBER OF CONTROLS | MAF<sup>a</sup> IN CONTROLS | P |
| --- | --- | --- | --- | --- | --- | --- |
| FTLD-TDP A | *TBK1* | 3 | 7.77X10<sup>-03</sup> | 0 | 0 | 1.27X10<sup>-11</sup> |
| FTLD-TDP B | *TBK1* | 5 | 8.68X10<sup>-03</sup> | 0 | 0 | 3.17X10<sup>-12</sup> |
| FTLD-TDP B | *VIPR1* | 3 | 5.21X10<sup>-03</sup> | 1 | 1.59X10<sup>-04</sup> | 4.65X10<sup>-7</sup> |
| FTLD-TDP C | *RBPJL* | 5 | 5.35X10<sup>-03</sup> | 3 | 4.76X10<sup>-04</sup> | 6.39X10<sup>-7</sup> |
| FTLD-TDP C | *L3MBTL1* | 8 | 8.57X10<sup>-03</sup> | 3 | 4.76X10<sup>-04</sup> | 2.38X10<sup>-7</sup> |

<sup>a</sup>MAF = minor allele frequency

## Discussion

In this work, we report 8 new genome-wide significant FTLD-TDP risk loci and 3 new genes harboring rare variants contributing to FTLD-TDP risk, by performing the largest FTLD-TDP WGS study to date, including 985 patients and 3,153 controls. A comprehensive analysis of our data highlights the genetic overlap between FTLD-TDP, ADRD, and ALS while also defining tissue and cell type enrichment unique to FTLD-TDP. Most importantly, we highlight distinct genetic aetiologies for each of the three main FTLD-TDP pathological subtypes (A, B and C) suggesting that multiple distinct pathomechanisms underlie the TDP-43 dysfunction and deposition in FTLD-TDP (**Fig. 7**).

**Figure 7:**
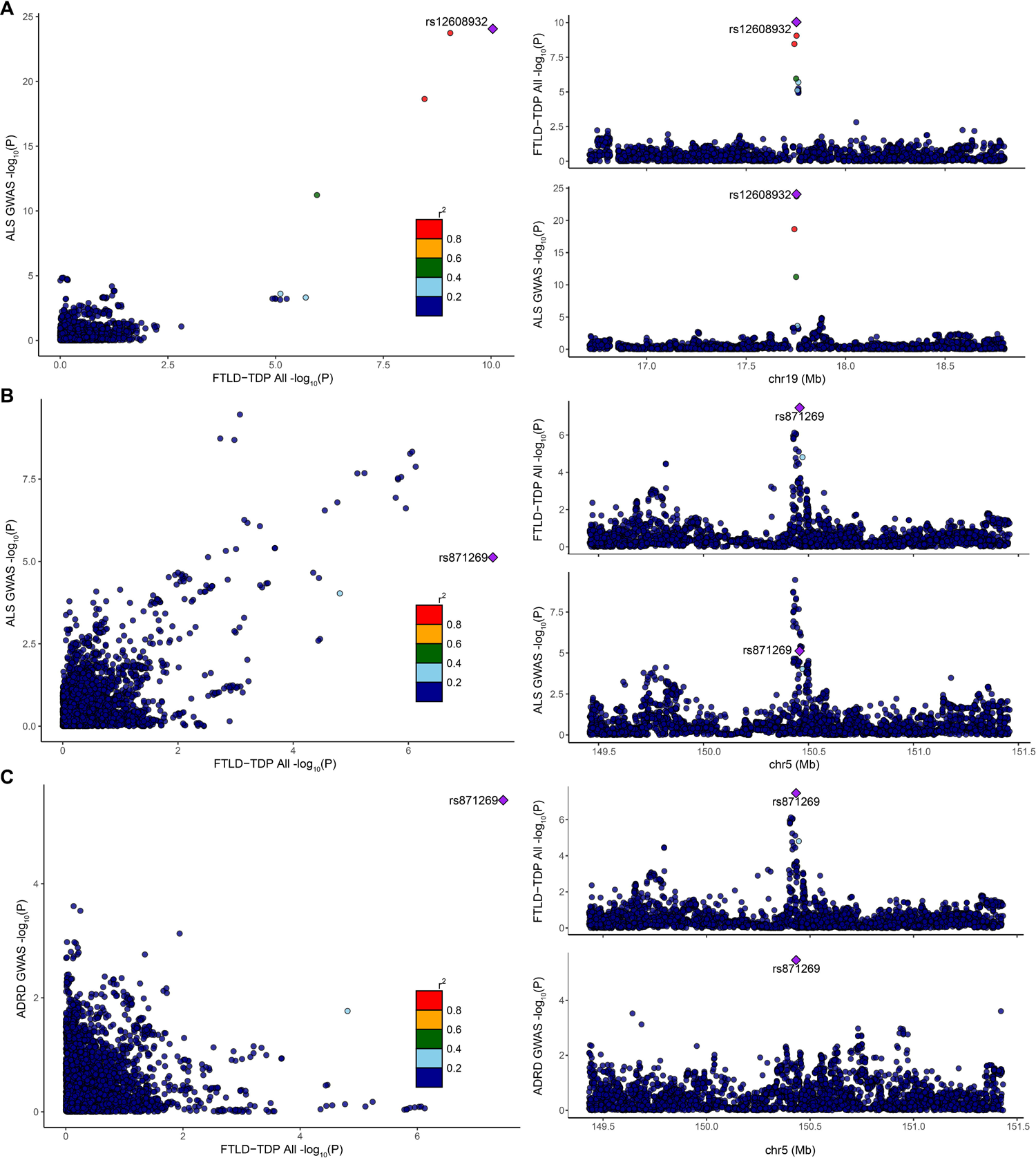
Schematic representation of findings from the International FTLD-TDP WGS phase II. Genome-wide significant single variant loci, exome-wide significant genes, enriched gene ontology pathways and tissues- and cell-types enriched for genome-wide significant risk loci are shown for each FTLD-TDP pathological subtype in rings moving from the center (genome-wide significant single variant loci in FTLD-TDP All) to the outer rings. Orange background shades correspond to FTLD-TDP A findings, green background shades to FTLD-TDP B findings and blue background shades to FTLD-TDP C findings. Gene names in green font were exome-wide significant using a gene-based approach with common variants while gene names in red font were exome-wide significant using a gene-based approach with rare variants. In addition to unique associations, some overlap between FTLD-TDP A and B exist (TBK1, lysosomal function and inflammatory response), whereas FTLD-TDP C showed unique and non-overlapping genetic profile.

We confirm and replicate for the first time our previously reported GWAS signal at the *UNC13A* locus in FTLD-TDP patients^11^. The *UNC13A* risk haplotype tagged by rs12973192 and rs12608932 was previously shown to increase cryptic splicing of *UNC13A* in brain tissue by modulating TDP-43 binding^18,19^. The cryptic splicing leads to transcripts with premature stop codons and the subsequent loss of UNC13A protein, significantly impacting the release of vesicles in glutamatergic synapses^20^. Since *UNC13A* thus represents a shared risk factor between ALS and FTLD-TDP additional genetic or environmental mechanisms likely drive disease presentation.

We further establish and replicate a novel genetic association between the *TNIP1* locus and FTLD-TDP. Recently, Restuadi *et al.* deeply characterized the *GPX3*/*TNIP1* locus associated with ALS and suggested that *GPX3* should be prioritized for deeper exploration into disease mechanisms related to this region^21^. *GPX3*, encoding for glutathione peroxidase 3, is a secreted enzyme involved in the regulation of oxidative damage, and its levels were found to be reduced in ALS sera^22^. Interestingly, however, the risk variant associated with FTLD-TDP (rs871269) is an expression quantitative trait locus for *TNIP1* in the dorsolateral prefrontal cortex, and along with the fact that we only observed a weak colocalization signal with the ALS locus, we highlight *TNIP1* as the most likely gene candidate for FTLD-TDP. In fact, we observed a shared signal at this locus between our FTLD-TDP GWAS and the recent large ADRD GWAS^14^, suggesting genetic and/or clinical overlap between AD and FTLD-TDP. TNIP1 is a ubiquitin-binding adaptor protein that regulates cell death and innate immune response through NF-kb activation^23–25^. TNIP1 undergoes phosphorylation by TBK1 and interacts with OPTN^26^, two proteins associated with FTLD-TDP etiology^12,27^. While this functional connection further supports *TNIP1* as FTLD-TDP risk gene, more work is needed to understand the mechanisms underlying disease onset. Overall, we substantiate the genetic overlap between ALS, ADRD, and FTLD-TDP and emphasize the need for deeper exploration into pathways underlying disease-specific risk.

One of the most striking conclusions from this phase II FTLD-TDP GWAS is the distinct association signals among FTLD-TDP pathological subtypes. Even the *UNC13A* and *TNIP1* risk loci, which reach genome-wide significance in the meta-analysis stage, show stronger association in FTLD-TDP B alone, and for the first time, genome-wide significant common risk loci are reported for each of the individual pathological FTLD-TDP subtypes.

In FTLD-TDP A, in addition to individual genome-wide significant common variants assigned to *GRN*, *TINAG*, *MZT1,* and *FARP2* risk loci, we identified exome-wide significant association with the burden of common variants in *GRN* and *TMEM106B*, in addition to multiple QTL-based analyses prioritizing *TMEM106B* as a tier 1 risk gene, re-enforcing the specific connection of these genes with FTLD-TDP A, even in patients without loss-of-function *GRN* mutations^11,28^. While *GRN* and *TMEM106B* are also reported as AD risk genes^14^, an even stronger connection exists between these genes and limbic-predominant age-related TDP-43 encephalopathy (LATE)^29,30^, which has a more restricted neuroanatomical distribution of TDP-43 pathology as compared to FTLD-TDP but with some characteristics of FTLD-TDP type A^31,32^. The *TMEM106B* signal is primarily influenced by rs10281425, a variant located in the 3’UTR of *TMEM106B,* which tags the previously reported *TMEM106B* risk haplotype^33^ associated with an increase in *TMEM106B* mRNA expression^33^ and a higher burden of insoluble disease-associated TMEM106B C-terminal fragments^34^. More broadly, also including prioritized genes from the subthreshold regions, gene ontology analysis in FTLD-TDP A revealed enrichment in genes implicated in lysosomal function driven by *GRN*, *TMEM106B* but also *CSTB*, three genes which also had the highest individual gene scores in the prioritization analysis in FTLD-TDP A. *CSTB* encodes one of the most abundant lysosomal proteases in the brain^35^, and has been reported as a progranulin protease^36,37^. Genes involved in lysosomal dysfunction were also overrepresented in FTLD-TDP B, including *GRN* and *PPT1*. PPT1, is a lysosomal enzyme which facilitates the degradation of fatty-acylated proteins by lysosomal hydrolases. Mutations in *PPT1* cause neuronal ceroid lipofuscinosis 1^38,39^ and *Ppt1* knock-out mice displayed fewer lipid droplets (LD) than wild type, indicating impairment of lipophagy, previously associated with FTLD/ALS^40–43^. Overall, our genetic data provides compelling evidence that lysosomal dysfunction contributes to the pathobiology of FTLD-TDP A, and, to a lesser extent FTLD-TDP B.

For FTLD-TDP B, additionally, we identified individual genome-wide significant associations with variants in the *RCL1* and *PDS5* loci, and we observed enrichment for gene ontology terms related to retrograde transport resulting from the *VPS53* and *DENND2A* loci. VPS53 is part of the Golgi-associated retrograde protein (GARP) complex^44,45^ involved in intracellular cholesterol transport by targeting NPC2 to lysosomes^46^. Recently, laser capture microdissection and single-cell mass spectrometry-based proteomics in motor neurons of ALS patients revealed a strong reduction in endolysosomal trafficking complexes such as the GARP complexes^47^. Limited information about DENN2A function is currently available, but structural and functional analysis indicates it may be involved in intracellular vesicle trafficking to the lysosome and to the Golgi through its guanine nucleotide exchange factor activity and regulation of RAB family GTPases^48^. However, retrograde transport has been previously implicated in ALS with, for instance, mutations in *DCTN1*^49,50^ and *KIF5A*^51,52^, highlighting functional connections of prioritized genes from the subthreshold loci with TDP-43 dysfunction and ALS. Future GWAS with larger sample sizes, potentially combining FTLD-TDP B and ALS, are required to firmly establish a genetic contribution of this pathway to disease.

Focusing on rare variants, exome-wide significant association with *TBK1* was observed in both FTLD-TDP A and B (but not FTLD-TDP C), confirming *TBK1* mutations as the most common cause of FTLD-TDP after *GRN* and *C9orf72*^12^. We further unveiled rare predicted pathogenic variants associated with FTLD-TDP B within *VIPR1,* which encodes for the vasoactive intestinal peptide receptor 1. The variants are predicted to lead to an alteration of VIPR1 function, impairing the vasoactive intestinal peptide (VIP) biological pathway. Indeed, VIPR1 is activated upon binding by VIP, which exerts a neuroprotective effect mainly through glial cells^53,54^ even though neurons also express VIPRs^55,56^. Notably, VIP is also a key regulator of innate and adaptive immunity^57^, making it an important therapeutic target for multiple neurodegenerative diseases. Altogether, our studies suggest that lysosome dysfunction and/or alterations in the innate and adaptive immune system are important contributors to both FTLD-TDP A and B risk, yet to varying degrees in each pathological subtype and with likely important variability in the contribution from each pathway among individual patients.

FTLD-TDP C was previously recognized as a clinicopathological entity distinct from FTLD-TDP A and B^58^, and our genetic studies support this notion showing no overlap in common or rare risk genes with the other FTLD-TDP types. Importantly, however, while often considered a sporadic FTLD subtype^13,59,60^, we implicate several genes and risk loci in FTLD-TDP C and uncover a potential role for mitochondrial membrane dysfunction and the notch signaling pathway. *C19orf52* (*TIMM29*), which mediates the import and insertion of multi-pass transmembrane proteins into the mitochondrial inner membrane, was identified as the first genome-wide significant risk locus for FTLD-TDP C. Rare-variant burden analyses further associated rare predicted pathogenic variants in the *RBPJL* gene with FTLD-TDP C. *RBPJL* encodes for the recombination signal binding protein for immunoglobulin kappa J region like transcription factor. RBPJL is able to repress Notch target gene expression (Hey1, Hey2, HeyL and Notch3)^61^. As such, our findings align with a previous analysis of sub-genome-wide significant genes in clinical svPPA patients, enriched in FTLD-TDP C, which highlighted an overrepresentation of the Notch pathway^62^. Interestingly *RBPJL* and *L3MBTL1*, the second gene carrying rare predicted pathogenic variants in FTLD-TDP C, are part of the same co-expression module, suggesting that they are functionally related. Moreover, L3MBTL1, a histone methyl-lysine binding protein, is a key regulator of proteotoxicity associated with *C9orf72* dipeptide repeats and mutant *SOD1*^63^ and was found to be increased in spinal cord of ALS patients. Furthermore, reduction of L3MBTL1 expression in drosophila models with the C9orf72-associated dipeptides poly(PR) or poly(GR) ameliorated the rough-eye phenotype^63^, suggesting that loss of L3MBTL1 expression is beneficial. While no RNA samples were accessible from rare variant carriers, nonsense-mediated decay escape has been reported in other genes linked to ALS^64^. It is thus possible that the *L3MBTL1* variants lead to the generation of truncated proteins with toxic gain-of-function, but additional work is necessary to understand the disease etiology fully.

When analyzed in sum, common variants associated with the different FTLD-TDP pathological subtypes appeared to be located in genes expressed in excitatory neurons in contrast to AD risk variants, which are enriched in microglia. Interestingly, glutamatergic transmission impairment has been reported in FTLD^65–69^ and voxel-based brain changes have been significantly associated with spatial distribution of mGluR5 in symptomatic *C9orf72* and *GRN* carriers^70^. Therefore, and in line with previously reported studies, our data suggest that neurons are the major players in disease etiology, as compared to what has been observed in ADRD. Interestingly, the distribution of risk loci was specific to the cerebellar hemisphere and the frontal-cortex for both FTLD-TDP A and B, as opposed to FTLD-TDP C where genes expressed in small intestine were enriched in risk loci. While the link between gut microbiome and FTLD remains limited^71^, our data suggest that the gut-brain axis might be of interest for future studies. In fact, emerging evidence also supports a role for the gut-brain axis in autoimmune diseases^72^, a group of disorders that were found to be enriched in svPPA patients^73^.

In prior studies, besides *UNC13A*, common variants in the *HLA* and *DPP6* loci were reported to be associated with FTLD^11,13^. The *HLA-DR5* locus was identified as associated with clinical FTLD but was not replicated in phase I of our International FTLD-TDP WGS Consortium. HLA-*DQA2* and *DPP6* loci were reported as overall FTLD-TDP risk loci in phase I^11^ but were not replicated in the current study. The relative composition of patients with FTLD-TDP pathological subtypes in phase I and II (e.g., less FTLD-TDP A in phase II) and inclusion of clinically diagnosed individuals in phase II may have contributed to this; however, it is also possible that the increase in sample size reduced type I errors from phase I. Importantly, we did identify and replicate in two independent cohorts the *UNC13A* and *TNIP1* loci associated with FTLD-TDP. Replication of the newly identified risk loci, each specific to distinct neuropathological FTLD-TDP subtypes, will require additional GWAS studies in the future. Obtaining sufficient samples will, however, be challenging, especially for FTLD-TDP A, which lacks a clear clinical correlate of the pathological phenotype. Functional characterization of the newly identified genes and loci may also provide mechanistic insight.

In conclusion, we confirmed *UNC13A* and identified 8 new genetic loci, i.e. *TNIP1*, *GRN*, *TINAG*, *MZT1*, *FARP2*, *RCL1*, *PDS5B,* and *C19orf52*, and 3 new genes with rare variants associated with FTLD-TDP risk, i.e. *VIPR1*, *RBPJL*, and *L3MBTL1*. By enriching in neuropathologically confirmed patients and substantially increasing our cohort size, we gained important knowledge in our understanding of FTLD-TDP pathophysiology. The recognition that individual FTLD-TDP subtypes could in fact be considered separate diseases with distinct pathomechanisms has significant implications for the design of clinical trials and therapeutic interventions.

## Methods

### Samples

Our current dataset includes previously generated data through the International FTLD-TDP WGS consortium phase I^11^ with 554 persons with clinicopathologically defined FTLD-TDP and newly generated phase II sequencing data from 32 FTLD-TDP A, 43 FTLD-TDP B, 66 FTLD-TDP C, 4 FTLD-TDP E, and 9 with unclassifiable FTLD-TDP pathology (abbreviated as FTLD-TDP U, **Supplementary Table 19**). To increase statistical power, we also sequenced 70 persons with clinical diagnosis of bvFTD/ALS, a clinical subtype associated with FTLD-TDP B, and 283 persons with svPPA, a clinical subtype associated with FTLD-TDP C. Overall, the total cohort pre-quality control was a combined FTLD-TDP cohort of 202 FTLD-TDP A, 237 FTLD-TDP B, 225 FTLD-TDP C, 4 FTLD-TDP D, 11 FTLD-TDP E, 29 FTLD-TDP U persons, 70 persons with bvFTD/ALS and 283 persons with svPPA. All persons clinically or pathologically diagnosed with FTLD are referred to as patients throughout the manuscript. We further used WGS data from 982 participants from the Mayo Clinic Biobank (from phase I)^11,74^, 322 new controls free of neurodegenerative disorder from Mayo Clinic with WGS available, and 2,037 controls derived from the Alzheimer’s disease sequencing project (ADSP). *C9orf72* repeat expansions were assessed in all patients using our previously reported two-step protocol and Sanger sequencing was used to perform mutation analyses of *GRN*^7,9^. Patients with *C9orf72* repeat expansion and LOF mutations in *GRN* were removed prior to WGS. Study protocols were reviewed and approved by the appropriate institutional review boards.

### Whole genome sequencing

In phase I of the International FTD-TDP WGS consortium, WGS was generated on 554 patients with FTLD-TDP (512 passed QC in that study)^11^. Briefly, whole blood- or brain-derived DNA from 499 unrelated FTLD-TDP patients and 982 participants from the Mayo Clinic Biobank Study were sequenced at HudsonAlpha using the standard library preparation protocol using NEBNext® DNA Library Prep Master Mix Set for Illumina® (New England BioLabs Inc., Ipswich, MA, USA), Concentration of the libraries was assessed by Qubit® 2.0 Fluorometer, and the quality of the libraries was estimated by a DNA 5 K chip on a Caliper GX. Accurate quantification was determined using the qPCR-based KAPA Biosystems Library Quantification kit (Kapa Biosystems, Inc., Woburn, MA, USA). Each sample was sequenced on one lane of Illumina’s HiSeq X instrument using v2 flow cells and reagents to target 30× genomic coverage. Fastq files previously generated on an Illumina HiSeq X for 55 FTLD-TDP patients were obtained from 3 sites: UCSF (n = 36), DZNE (n = 14) and NSW (n = 5). In phase II, additional WGS of 507 patients with FTLD-TDP, svPPA, bvFTD/ALS and 322 controls free of neurodegenerative disorders was performed at USUHS sequencing center or Mayo Clinic Rochester using the TruSeq DNA PCR-Free Library preparation Kit (Illumina) followed by Whole Genome Sequencing by synthesis (SBS) chemistry on HiSeq X Illumina platform using the HiSeq X Ten Reag. kit v2.5. For all patients and controls, fastq files were transferred to Mayo Clinic and processed through the Mayo Genome GPS v4.0 pipeline in batches of up to 75 samples. Briefly, reads were mapped to the human reference sequence (GRCh38 build) using the Burrows–Wheeler Aligner, and local realignment around indels was performed using the Genome Analysis Toolkit (GATK). Variant calling was performed using GATK HaplotypeCaller followed by variant recalibration (VQSR) according to the GATK best practice recommendations. At the time of analysis, participants from the Mayo Clinic Biobank with possible clinical diagnosis or family history of a neurodegenerative disorder were removed. We also included genotypic data (vcf) from 2,037 controls from the Alzheimer’s Disease Sequencing Project, leading to a total of 1,061 patients and 3,341 controls.

### Sample level quality control and definition of subgroups

Samples with less than 30× coverage in more than 50% of the genome, call rate below 85%, sex error, contamination defined by a FREEMIX score above 4 or non-Caucasian ethnicity were removed. At this step, joint genotyping on all samples was performed, a final relatedness measurement was calculated using PREST^75^, and duplicates were removed, while only one individual per family was kept. In total, 985 pathologically confirmed FTLD-TDP or presumed FTLD-TDP patients clinically presenting with svPPA or bvFTD/ALS, as well as 3,153 neurologically normal controls passed all quality control measures (**Supplementary Table 1**). Age at onset of svPPA and bvFTD/ALS did not differ from the age at onset of FTLD-TDP C (*P*=1) and FTLD-TDP B patients (*P*=1), respectively. Based on these findings and the previously established associations between the svPPA and bvFTD/ALS clinical diagnoses with specific FTLD-TDP pathological subtypes, we combined svPPA with FTLD-TDP C and bvFTD/ALS with FTLD-TDP B patients in all analyses. Within our overall cohort of 193 FTLD-TDP A, 288 FTLD-TDP B (defined as FTLD-TDP B and bvFTD/ALS) and 467 FTLD-TDP C (defined as FTLD-TDP C and svPPA), the ages at onset and death differed significantly between the pathological FTLD-TDP subtypes (**Table 2; Supplementary** Figure 1). FTLD-TDP A patients had a later age at onset than FTLD-TDP B and FTLD-TDP C groups (*P*=4.73×10^-8^, *P*=1.37×10^-13^, respectively), and a later age at death (*P*=4.00×10^-15^, *P*=3.00×10^-6^, respectively). FTLD-TDP B had an earlier age at death as compared to FTLD-TDP C (*P*=5.60×10^-8^). Differences in age distribution in between patient groups were assessed using the Kruskal-Wallis test followed by Wilcoxon test correcting for multiple testing. Corrected *P* are provided.

### Variant level quality control

Genotype calls with genotype quality (GQ) < 20 and/or depth (DP) < 10 were set to missing, and variants with edit-distance > 4 and call rate < 80% were removed from all subsequent analyses leading to a total of 85,345,466 variants. For all analyses, only variants that pass VQSR (127,658 variants removed) and with a call rate > 95% in cases or controls were considered (591,431 variants removed). Functional annotation of variants was performed using ANNOVAR (version2016Feb01). Rare loss of function variants (frameshift insertion/deletion/block substitution, stopgain, stoploss and splicing single nucleotide variants—SNVs) identified in exome-wide significant associated genes in the rare variant analyses (**Supplementary Table 17**) were confirmed in patients by Sanger sequencing (primers available upon request). For the known neurodegenerative disease genes (*GRN*, *MAPT*, *TBK1*, *OPTN*, *VCP*, *TARDBP*, *CHCHD10*, *SQSTM1*, *UBQLN2*, *hnRNPA1*, *hnRNPA2B1*, *CSF1R*, *FUS*, *CHMP2B*, and *LRRK2),* potentially pathogenic rare variants were also identified and confirmed by Sanger sequencing (n=25; **Supplementary Table 20**).

### Generation of principal components

Prior to running genetic association analyses, principal component (PC) analysis was performed using a subset of variants meeting the following criteria: minor allele frequency (MAF)>5% and full sample Hardy–Weinberg Equilibrium (HWE) *P*>1×10^-5^. Influential regions such as the *HLA* region were removed, and variants were pruned by linkage disequilibrium with r^2^ threshold of 0.1 prior to PC analysis. This analysis identified 13 PCs that were significantly associated with patient/control status, which were subsequently used as covariates in all genetic association analyses.

### Variant-level analysis of common variants

For the common variant GWAS, single nucleotide variants (SNV) with MAF>0.01 in patients or controls (n=7,178,250 variants) and HWE *P*>1.00×10^-6^ in controls were analyzed (17,450 variants removed). In addition, since whole genome sequencing of FTLD-TDP patients and controls was performed at multiple sites, a test was performed to identify variants with significant differences in genotype distributions between sequencing batches, and 592,701 SNVs showing evidence of batch effects (p < 0.05) were removed leading to a total of 6,568,099 variants analyzed.

For all remaining variants, association of genotypes with the patient/control status was assessed using logistic regression with allele dosage as the predictor assuming log-additive allele effects. Sex and the first thirteen PCs were included as covariates in the models. The SNV-level analyses were performed using PLINKv.00a23LM2 combining all FTLD-TDP cases (FTLD-TDP All) and in FTLD-TDP pathological subtypes. Meta-analyses of FTLD-TDP phase II with publicly available dataset from the Dementia-seq project (phs001963.v2.p1) was performed under a fixed-effects model comparing our data with 2,102 FTLD cases and 1,748 controls from the Dementia-seq project using Metal^76^. Dementia-seq vcf were processed the exact same way as our data except that 10 PCs were included in the model to perform common variant association analysis.

### Colocalization analyses

We performed colocalization analysis for *UNC13A* and *TNIP1* loci (top SNVs ±100 kb) with ALS (GCST90027164) and ADRD (GCST90027158) using the ‘coloc’ package version 4.0.4 in R using our meta-analyses data. When the summary statistics of the other trait was expressed on another build than GRCh38, the variant alleles and positions were converted. We set the prior probabilities to π1 = 1×10^−4^, π2 = 1×10^−4^ and π12 = 1×10^−5^ for a causal variant in trait 1 or trait 2 and a shared causal variant between traits 1 and 2, respectively (default parameters). Sensitivity analysis was performed at π12=1×10^-6^. *P* <0.05 was considered statistically significant.

### Tissue and cell type enrichment analysis

Tissue and cell type enrichment analyses were performed using the summary statistics and FUMA. Briefly, FUMA aggregates summary statistics per gene to calculate gene-wise association signals using MAGMA version 1.6 and subsequently tests whether tissues and cell types are enriched for expression of these genes. For tissue enrichment analysis, we used the GTEx version 8 reference set. *P*<0.05 across all tissues (n = 54) were considered statistically significant. For cell type enrichment analyses, we used human-derived single-cell RNA-seq data from major brain cell types (PsychENCODE). Excitatory and inhibitory neurons from the PsychENCODE dataset were labeled based on their transcriptional profile from 1 to 8^77^. *P*<0.05 were considered statistically significant.

### Gene prioritization and functional interpretation of GWAS

We performed the gene prioritization and functional interpretation analyses for FTLD-TDP All and each FTLD-TDP pathological subtype separately by using the subtype-specific GWAS summary statistics. We adapted a systematic gene prioritization and functional interpretation strategy (as previously described in Bellenguez et al.^14^) to prioritize GWAS-implicated candidate risk genes and nominate possible downstream biological mechanisms. Briefly, six distinct domains, that are related to lead variant annotation and molecular QTL-GWAS integration analyses (e.g., colocalization and TWAS) in FTLD-relevant tissues and cell types were systematically assessed: (1) variant annotation, (2) eQTL-GWAS integration, (3) sQTL-GWAS integration, (4) protein expression QTL (pQTL)-GWAS integration, (5) mQTL-GWAS integration, and (6) histone acetylation QTL (haQTL)-GWAS integration; for which detailed information on categories and subcategories is provided in **Supplementary Table 2.**

In the variant annotation domain, for each lead variant at each locus, we queried which candidate risk genes were the nearest protein-coding genes with respect to the genomic position of the lead variants, and/or whether the lead variant was a rare (MAF < 1% in gnomAD v4 non-Finnish European samples) and/or protein-altering (missense or predicted LOF) variant for the same nearest protein-coding genes. In the molecular QTL-GWAS integration domains, we leveraged molecular cis-QTL catalogues for different molecular phenotypes (i.e., gene expression, splicing, protein expression, methylation, and histone acetylation) in FTLD-relevant tissues and cell types, we performed genetic colocalization analyses between molecular cis-QTL and GWAS signals, TWAS, and proteome-wide association studies (PWAS). For these analyses, we processed and used publicly available molecular QTL catalogues; namely, FTLD-relevant bulk brain regions from AMP-AD^78–81^ (as reanalyzed in Bellenguez et al.^14^) and GTEx v8^82^ cohorts for the bulk brain eQTLs and sQTLs, eight major brain cell types (excitatory neurons, inhibitory neurons, astrocytes, oligodendrocytes, microglia, oligodendrocyte precursor cells/committed oligodendrocyte precursors [OPCs/COPs], pericytes, and endothelial cells) from Bryois et al.^83^ and primary microglia from Young et al.^84^ and the MiGA study^85^ for the brain cell-type-specific eQTLs (ct-eQTL) and for microglia sQTLs (from the MiGA study), dorsolateral prefrontal cortex (DLPFC) pQTLs from Wingo et al.^86^ (v2), and DLPFC mQTLs and haQTLs from Brain xQTL serve (June 2021 release)^86,87^. Finally, we also included naïve state monocyte and macrophage eQTL catalogues^88–93^ reanalyzed by eQTL Catalogue (Release 6)^94^ and lymphoblastoid cell line (LCL) eQTLs from GTEx v8^82^ and the European Alzheimer & Dementia Biobank (EADB) Belgian LCL cohorts^14^. Using each of these molecular QTL catalogues, we first investigated whether the reported lead variants in this study were significant molecular QTLs for the quantified levels of molecular phenotypes in tissues and cell types of interest. Moreover, for each quantified molecular phenotype in these catalogues we performed molecular QTL-GWAS coloc (v5.2.2) analyses to determine if specific molecular QTL signals are colocalized (at coloc PP4 ≥ 70%) with FTLD subtype GWAS signals. Finally, we conducted TWAS (using FUSION and S-PrediXcan [implemented in MetaXcan] tools) for each heritable feature modelled in gene expression (eTWAS; followed by eTWAS fine mapping with FOCUS^95^ [v0.803] within 1 Mb extended genome-wide significant lead variant genetic regions in each FTLD-TDP subtype GWAS), splicing (sTWAS), and PWAS reference panels derived from AMP-AD bulk brain^78–81^, GTEx bulk brain and LCL^82^, EADB Belgian LCL^14^, and Wingo et al. DLPFC data^86^, to identify the significant associations (after Bonferroni correction) between predicted levels of gene expression, splicing, and protein expression with each FTLD subtype-specific genetic risk. Detailed description and details (e.g., number of samples, significance criteria, references and sources) of these molecular QTL catalogues used in this study for the systematic gene prioritization strategy and functional interpretation of FTLD-TDP GWAS results can be found in **Supplementary Table 5**.

Using a predetermined weighting scheme for each type of evidence (see **Supplementary Table 3**), we computed a gene prioritization score (between 0 and 87) for each gene which was constructed by the weighted sum of the hits in different subcategories within six distinct domains described above. As described in Bellenguez et al.^14^ in detail, we gave higher weights for the hits obtained through the brain QTLs rather than other tissue QTLs, for the replicated hits across multiple catalogues or reference panels, and for the fine-mapped eTWAS hits. After obtaining weighted gene prioritization scores in each FTLD-TDP subtype-specific gene prioritization analysis, we first assigned each candidate risk gene (with gene prioritization score >0) to the genome-wide significant loci if their gene coordinates (based on GENCODE v24) are positioned within a ±1 Mb window of the identified lead variants (**Table 2**). The rest of the candidate risk genes in subthreshold regions (nominated by coloc and TWAS analyses only) were grouped together if they were positioned together (<1 Mb), and these subthreshold regions were indexed and named as subthreshold loci. The candidate risk genes in genome-wide significant and subthreshold loci were also annotated by the evidence of minimum *P* observed within 1 Mb of the gene coordinates in related FTLD-TDP subtype-specific GWAS summary statistics. We then ranked the protein-coding genes per locus in each FTLD subtype-specific analysis based on their total weighted scores, and investigated the relative score differences between the highest ranked protein-coding gene and the other candidate risk genes in each locus, together with the overall total weighted score of the top-ranked gene. We then classified candidate risk genes in each locus as tier 1 and tier 2 prioritized risk genes, respectively having a higher and lower level of confidence for being a true risk gene in a given locus (see Bellenguez et al.^14^ for detailed description). As also described in Bellenguez et al.^14^, the gene prioritization pipeline determines a single tier 1 prioritized risk gene in each locus if there is adequate evidence, meanwhile additional tier 2 prioritized risk genes in the same loci or multiple tier 2 prioritized risk genes in a locus can also be assigned based on the score distribution of candidate genes in the investigated loci.

### Gene Ontology analyses

Gene ontology on tier 1 genes identified in FTLD-TDP All or in individuals FTLD-TDP subtype analyses were performed using anRichment R package which aggregates summary statistics and assesses gene ontology term enrichment. Gene ontology terms were collapsed using the rrvigo R package. Only terms with 2 or more genes were considered in the analyses. *P* <0.05 was considered statistically significant.

### Gene-level analysis of rare variants

Association of rare variants with the patient/control status was assessed using an unweighted burden test implemented using the SKAT_1.2.1 R package. Only VQSR pass variants with call rate > 90%, ED ≤ 4, and MAF < 0.01 in either patients or controls were included. We included only frameshift (insertion/deletion/block substitution), stopgain, stoploss and splicing SNVs (jointly defined as loss-of-function (LOF) variants), and non-synonymous SNVs with REVEL score above 0.75^96^. Sex and the first thirteen PCs were used as covariates. Genome-wide significance was defined as *P*<5×10^-8^ and exome-wide significance as a p value < 2.5×10^-6^ (Bonferroni correction for 20,000 genes).

### *RBPJL* and *L3MBTL1* RNA expression

Assessment of module membership of *RBPJL* and *L3MBTL1* was performed using the gene co-expression analysis from the BrainEXP-NPD^17^ website using default parameters. Single nuclei RNA expression was assessed using the transcriptomic comparative viewer if the Seattle Alzheimer’s Disease Brain cell Atlas from middle temporal gyrus of 84 aged donors (42 cognitively normal and 42 with dementia).

## Data Availability

Summary statistics will be available on dbGAP platform post publication. Datasets and molecular QTLs used in the gene prioritization are publicly available (see also Supplementary Table 4): eQTLs and eTWAS reference panels in AD-relevant bulk brain regions from AMP-AD cohorts and in LCLs from the EADB Belgian cohort, as analyzed by Bellenguez et al.^14^: https://doi.org/10.5281/zenodo.5745927; sQTLs and sTWAS reference panels in AD-relevant bulk brain regions from AMP-AD cohorts and in LCLs from the EADB Belgian cohort, as analyzed by Bellenguez et al^14^.: https://doi.org/10.5281/zenodo.5745929; Bryois et al.^83^ ct-eQTL catalogues (https://doi.org/10.5281/zenodo.5543734); eQTL Catalogue database (https://www.ebi.ac.uk/eqtl/); Brain xQTL serve mQTL and haQTL catalogues (https://mostafavilab.stat.ubc.ca/xqtl/xQTL_updated_data/); GTEx v8 eQTL and sQTL catalogues (https://www.gtexportal.org/); GTEx v8 expression and splicing prediction models for eTWAS/sTWAS (https://predictdb.org/post/2021/07/21/gtex-v8-models-on-eqtl-and-sqtl/#mashr-based-models); MiGA eQTLs (https://doi.org/10.5281/zenodo.4118605); MiGA sQTLs (https://doi.org/10.5281/zenodo.4118403); MiGA meta-analysis (https://doi.org/10.5281/zenodo.4118676); and Wingo et al.^86^ pQTLs v2 (https://www.synapse.org/#!Synapse:syn23627957).

## Declarations

### Ethics approval and consent to participate

This study was approved by the appropriate Mayo Clinic Institutional Review Board.

### Consent for publication

Not applicable.

## Acknowledgements

We thank the Mayo Clinic Center of Individualized Medicine for collection and sequencing of the Mayo Clinic Biobank samples. CG would also like to acknowledge Dr Inger Nennesmo for the neuropathological characterization. Acknowledgment for the ADSP dataset can be found in Supplementary Notes.

## References

1. Rascovsky, K. et al. Sensitivity of revised diagnostic criteria for the behavioural variant of frontotemporal dementia. Brain 134, 2456–77 (2011).

2. Gorno-Tempini, M.L. et al. Classification of primary progressive aphasia and its variants. Neurology 76, 1006–14 (2011).

3. Lee, E.B. et al. Expansion of the classification of FTLD-TDP: distinct pathology associated with rapidly progressive frontotemporal degeneration. Acta Neuropathol 134, 65–78 (2017).

4. Mackenzie, I.R. et al. A harmonized classification system for FTLD-TDP pathology. Acta Neuropathol 122, 111–3 (2011).

5. Mackenzie, I.R. et al. Heterogeneity of ubiquitin pathology in frontotemporal lobar degeneration: classification and relation to clinical phenotype. Acta Neuropathol 112, 539–49 (2006).

6. Mackenzie, I.R. & Neumann, M. Reappraisal of TDP-43 pathology in FTLD-U subtypes. Acta Neuropathol 134, 79–96 (2017).

7. Baker, M. et al. Mutations in progranulin cause tau-negative frontotemporal dementia linked to chromosome 17. Nature 442, 916–9 (2006).

8. Cruts, M. et al. Null mutations in progranulin cause ubiquitin-positive frontotemporal dementia linked to chromosome 17q21. Nature 442, 920–4 (2006).

9. DeJesus-Hernandez, M. et al. Expanded GGGGCC hexanucleotide repeat in noncoding region of C9ORF72 causes chromosome 9p-linked FTD and ALS. Neuron 72, 245–56 (2011).

10. Renton, A.E. et al. A hexanucleotide repeat expansion in C9ORF72 is the cause of chromosome 9p21-linked ALS-FTD. Neuron 72, 257–68 (2011).

11. Pottier, C. et al. Genome-wide analyses as part of the international FTLD-TDP whole-genome sequencing consortium reveals novel disease risk factors and increases support for immune dysfunction in FTLD. Acta Neuropathol 137, 879–899 (2019).

12. Pottier, C. et al. Whole-genome sequencing reveals important role for TBK1 and OPTN mutations in frontotemporal lobar degeneration without motor neuron disease. Acta Neuropathol 130, 77–92 (2015).

13. Ferrari, R. et al. Frontotemporal dementia and its subtypes: a genome-wide association study. Lancet Neurol 13, 686–99 (2014).

14. Bellenguez, C. et al. New insights into the genetic etiology of Alzheimer’s disease and related dementias. Nat Genet 54, 412–436 (2022).

15. Project Min, E.A.L.S.S.C. Project MinE: study design and pilot analyses of a large-scale whole-genome sequencing study in amyotrophic lateral sclerosis. Eur J Hum Genet 26, 1537–1546 (2018).

16. Benyamin, B. et al. Cross-ethnic meta-analysis identifies association of the GPX3-TNIP1 locus with amyotrophic lateral sclerosis. Nat Commun 8, 611 (2017).

17. Jiao, C. et al. BrainEXP: a database featuring with spatiotemporal expression variations and co-expression organizations in human brains. Bioinformatics 35, 172–174 (2019).

18. Brown, A.L. et al. TDP-43 loss and ALS-risk SNPs drive mis-splicing and depletion of UNC13A. Nature 603, 131–137 (2022).

19. Ma, X.R. et al. TDP-43 represses cryptic exon inclusion in the FTD-ALS gene UNC13A. Nature 603, 124–130 (2022).

20. Augustin, I., Rosenmund, C., Sudhof, T.C. & Brose, N. Munc13-1 is essential for fusion competence of glutamatergic synaptic vesicles. Nature 400, 457–61 (1999).

21. Restuadi, R. et al. Functional characterisation of the amyotrophic lateral sclerosis risk locus GPX3/TNIP1. Genome Med 14, 7 (2022).

22. Tanaka, H. et al. ITIH4 and Gpx3 are potential biomarkers for amyotrophic lateral sclerosis. J Neurol 260, 1782–97 (2013).

23. Oshima, S. et al. ABIN-1 is a ubiquitin sensor that restricts cell death and sustains embryonic development. Nature 457, 906–9 (2009).

24. Su, Z. et al. ABIN-1 heterozygosity sensitizes to innate immune response in both RIPK1-dependent and RIPK1-independent manner. Cell Death Differ 26, 1077–1088 (2019).

25. Zhou, J. et al. A20-binding inhibitor of NF-kappaB (ABIN1) controls Toll-like receptor-mediated CCAAT/enhancer-binding protein beta activation and protects from inflammatory disease. Proc Natl Acad Sci U S A 108, E998–1006 (2011).

26. Lee, Y. et al. Coordinate regulation of the senescent state by selective autophagy. Dev Cell 56, 1512–1525 e7 (2021).

27. Freischmidt, A. et al. Haploinsufficiency of TBK1 causes familial ALS and fronto-temporal dementia. Nat Neurosci 18, 631–6 (2015).

28. Mackenzie, I.R. et al. The neuropathology of frontotemporal lobar degeneration caused by mutations in the progranulin gene. Brain 129, 3081–90 (2006).

29. Katsumata, Y. et al. Multiple gene variants linked to Alzheimer’s-type clinical dementia via GWAS are also associated with non-Alzheimer’s neuropathologic entities. Neurobiol Dis 174, 105880 (2022).

30. Katsumata, Y., Nelson, P.T., Ellingson, S.R. & Fardo, D.W. Gene-based association study of genes linked to hippocampal sclerosis of aging neuropathology: GRN, TMEM106B, ABCC9, and KCNMB2. Neurobiol Aging 53, 193 e17-193 e25 (2017).

31. Nelson, P.T. et al. Limbic-predominant age-related TDP-43 encephalopathy (LATE): consensus working group report. Brain 142, 1503–1527 (2019).

32. Josephs, K.A. et al. Pathological, imaging and genetic characteristics support the existence of distinct TDP-43 types in non-FTLD brains. Acta Neuropathol 137, 227–238 (2019).

33. Van Deerlin, V.M. et al. Common variants at 7p21 are associated with frontotemporal lobar degeneration with TDP-43 inclusions. Nat Genet 42, 234–9 (2010).

34. C, T.V., et al. C-terminal TMEM106B fragments in human brain correlate with disease-associated TMEM106B haplotypes. Brain 146, 4055–4064 (2023).

35. Hook, V. et al. Cathepsin B in neurodegeneration of Alzheimer’s disease, traumatic brain injury, and related brain disorders. Biochim Biophys Acta Proteins Proteom 1868, 140428 (2020).

36. Lee, C.W. et al. The lysosomal protein cathepsin L is a progranulin protease. Mol Neurodegener 12, 55 (2017).

37. Zhou, X. et al. Lysosomal processing of progranulin. Mol Neurodegener 12, 62 (2017).

38. Vesa, J. et al. Mutations in the palmitoyl protein thioesterase gene causing infantile neuronal ceroid lipofuscinosis. Nature 376, 584–7 (1995).

39. Camp, L.A., Verkruyse, L.A., Afendis, S.J., Slaughter, C.A. & Hofmann, S.L. Molecular cloning and expression of palmitoyl-protein thioesterase. J Biol Chem 269, 23212–9 (1994).

40. Branchu, J. et al. Loss of spatacsin function alters lysosomal lipid clearance leading to upper and lower motor neuron degeneration. Neurobiol Dis 102, 21–37 (2017).

41. Han, S.M. et al. VAPB/ALS8 MSP ligands regulate striated muscle energy metabolism critical for adult survival in caenorhabditis elegans. PLoS Genet 9, e1003738 (2013).

42. Cingolani, F. & Czaja, M.J. Regulation and Functions of Autophagic Lipolysis. Trends Endocrinol Metab 27, 696–705 (2016).

43. Liu, Y. et al. A C9orf72-CARM1 axis regulates lipid metabolism under glucose starvation-induced nutrient stress. Genes Dev 32, 1380–1397 (2018).

44. Conibear, E. & Stevens, T.H. Vps52p, Vps53p, and Vps54p form a novel multisubunit complex required for protein sorting at the yeast late Golgi. Mol Biol Cell 11, 305–23 (2000).

45. Liewen, H. et al. Characterization of the human GARP (Golgi associated retrograde protein) complex. Exp Cell Res 306, 24–34 (2005).

46. Wei, J. et al. The GARP Complex Is Involved in Intracellular Cholesterol Transport via Targeting NPC2 to Lysosomes. Cell Rep 19, 2823–2835 (2017).

47. 47. Guise, A.J., et al. TDP-43-stratified single-cell proteomic profiling of postmortem human spinal motor neurons reveals protein dynamics in amyotrophic lateral sclerosis. *bioRxiv* (2023).

48. Yoshimura, S., Gerondopoulos, A., Linford, A., Rigden, D.J. & Barr, F.A. Family-wide characterization of the DENN domain Rab GDP-GTP exchange factors. J Cell Biol 191, 367–81 (2010).

49. Deshimaru, M. et al. DCTN1 Binds to TDP-43 and Regulates TDP-43 Aggregation. Int J Mol Sci 22(2021).

50. He, J., Yu, W., Liu, X. & Fan, D. An identical DCTN1 mutation in two Chinese siblings manifest as dHMN and ALS respectively: a case report. Amyotroph Lateral Scler Frontotemporal Degener 23, 149–153 (2022).

51. Nicolas, A. et al. Genome-wide Analyses Identify KIF5A as a Novel ALS Gene. Neuron 97, 1267–1288 (2018).

52. Xia, C.H. et al. Abnormal neurofilament transport caused by targeted disruption of neuronal kinesin heavy chain KIF5A. J Cell Biol 161, 55–66 (2003).

53. Brenneman, D.E., Hauser, J., Spong, C.Y. & Phillips, T.M. Chemokines released from astroglia by vasoactive intestinal peptide. Mechanism of neuroprotection from HIV envelope protein toxicity. Ann N Y Acad Sci 921, 109–14 (2000).

54. Brenneman, D.E. et al. Vasoactive intestinal peptide. Link between electrical activity and glia-mediated neurotrophism. Ann N Y Acad Sci 897, 17–26 (1999).

55. Chneiweiss, H., Glowinski, J. & Premont, J. Vasoactive intestinal polypeptide receptors linked to an adenylate cyclase, and their relationship with biogenic amine- and somatostatin-sensitive adenylate cyclases on central neuronal and glial cells in primary cultures. J Neurochem 44, 779–86 (1985).

56. Hosli, E. & Hosli, L. Autoradiographic localization of binding sites for vasoactive intestinal peptide and angiotensin II on neurons and astrocytes of cultured rat central nervous system. Neuroscience 31, 463–70 (1989).

57. Delgado, M. & Ganea, D. Vasoactive intestinal peptide: a neuropeptide with pleiotropic immune functions. Amino Acids 45, 25–39 (2013).

58. Baer, G.M. et al. Sporadic FTLD TDP type C is a distinct clinicopathological entity from FTLD types A/B (S7.007). Neurology 88, S7.007 (2017).

59. Rohrer, J.D. et al. The heritability and genetics of frontotemporal lobar degeneration. Neurology 73, 1451–6 (2009).

60. Snowden, J., Neary, D. & Mann, D. Frontotemporal lobar degeneration: clinical and pathological relationships. Acta Neuropathol 114, 31–8 (2007).

61. Pan, L., et al. Transcription Factor RBPJL Is Able to Repress Notch Target Gene Expression but Is Non-Responsive to Notch Activation. Cancers (Basel) 13(2021).

62. Bonham, L.W. et al. Genetic variation across RNA metabolism and cell death gene networks is implicated in the semantic variant of primary progressive aphasia. Sci Rep 9, 10854 (2019).

63. Lu, J. et al. L3MBTL1 regulates ALS/FTD-associated proteotoxicity and quality control. Nat Neurosci 22, 875–886 (2019).

64. Guissart, C. et al. Premature termination codons in SOD1 causing Amyotrophic Lateral Sclerosis are predicted to escape the nonsense-mediated mRNA decay. Sci Rep 10, 20738 (2020).

65. Murley, A.G. & Rowe, J.B. Neurotransmitter deficits from frontotemporal lobar degeneration. Brain 141, 1263–1285 (2018).

66. Ferrer, I. Neurons and their dendrites in frontotemporal dementia. Dement Geriatr Cogn Disord 10 **Suppl 1**, 55–60 (1999).

67. Premi, E. et al. Unravelling neurotransmitters impairment in primary progressive aphasias. Hum Brain Mapp 44, 2245–2253 (2023).

68. Bowen, D.M. et al. Imbalance of a serotonergic system in frontotemporal dementia: implication for pharmacotherapy. Psychopharmacology (Berl) 196, 603–10 (2008).

69. Procter, A.W., Qurne, M. & Francis, P.T. Neurochemical features of frontotemporal dementia. Dement Geriatr Cogn Disord 10 **Suppl 1**, 80–4 (1999).

70. Premi, E. et al. Early neurotransmitters changes in prodromal frontotemporal dementia: A GENFI study. Neurobiol Dis 179, 106068 (2023).

71. Ghezzi, L., Cantoni, C., Rotondo, E. & Galimberti, D. The Gut Microbiome-Brain Crosstalk in Neurodegenerative Diseases. Biomedicines 10(2022).

72. Khawar, M.M., Sr., et al. The Gut-Brain Axis in Autoimmune Diseases: Emerging Insights and Therapeutic Implications. Cureus 15, e48655 (2023).

73. Miller, Z.A. et al. TDP-43 frontotemporal lobar degeneration and autoimmune disease. J Neurol Neurosurg Psychiatry 84, 956–62 (2013).

74. Olson, J.E. et al. The Mayo Clinic Biobank: a building block for individualized medicine. Mayo Clin Proc 88, 952–62 (2013).

75. McPeek, M.S. & Sun, L. Statistical tests for detection of misspecified relationships by use of genome-screen data. Am J Hum Genet 66, 1076–94 (2000).

76. Willer, C.J., Li, Y. & Abecasis, G.R. METAL: fast and efficient meta-analysis of genomewide association scans. Bioinformatics 26, 2190–1 (2010).

77. Lake, B.B. et al. Integrative single-cell analysis of transcriptional and epigenetic states in the human adult brain. Nat Biotechnol 36, 70–80 (2018).

78. Allen, M. et al. Human whole genome genotype and transcriptome data for Alzheimer’s and other neurodegenerative diseases. Sci Data 3, 160089 (2016).

79. Mostafavi, S. et al. A molecular network of the aging human brain provides insights into the pathology and cognitive decline of Alzheimer’s disease. Nat Neurosci 21, 811–819 (2018).

80. Bennett, D.A. et al. Religious Orders Study and Rush Memory and Aging Project. J Alzheimers Dis 64, S161–S189 (2018).

81. Wang, M. et al. The Mount Sinai cohort of large-scale genomic, transcriptomic and proteomic data in Alzheimer’s disease. Sci Data 5, 180185 (2018).

82. Consortium, G.T. The GTEx Consortium atlas of genetic regulatory effects across human tissues. Science 369, 1318–1330 (2020).

83. Bryois, J. et al. Cell-type-specific cis-eQTLs in eight human brain cell types identify novel risk genes for psychiatric and neurological disorders. Nat Neurosci 25, 1104–1112 (2022).

84. Young, A.M.H. et al. A map of transcriptional heterogeneity and regulatory variation in human microglia. Nat Genet 53, 861–868 (2021).

85. Lopes, K.P. et al. Genetic analysis of the human microglial transcriptome across brain regions, aging and disease pathologies. Nat Genet 54, 4–17 (2022).

86. Wingo, A.P. et al. Integrating human brain proteomes with genome-wide association data implicates new proteins in Alzheimer’s disease pathogenesis. Nat Genet 53, 143–146 (2021).

87. Ng, B. et al. An xQTL map integrates the genetic architecture of the human brain’s transcriptome and epigenome. Nat Neurosci 20, 1418–1426 (2017).

88. Alasoo, K. et al. Shared genetic effects on chromatin and gene expression indicate a role for enhancer priming in immune response. Nat Genet 50, 424–431 (2018).

89. Nedelec, Y. et al. Genetic Ancestry and Natural Selection Drive Population Differences in Immune Responses to Pathogens. Cell 167, 657–669 e21 (2016).

90. Chen, L. et al. Genetic Drivers of Epigenetic and Transcriptional Variation in Human Immune Cells. Cell 167, 1398–1414 e24 (2016).

91. Momozawa, Y. et al. IBD risk loci are enriched in multigenic regulatory modules encompassing putative causative genes. Nat Commun 9, 2427 (2018).

92. Quach, H. et al. Genetic Adaptation and Neandertal Admixture Shaped the Immune System of Human Populations. Cell 167, 643–656 e17 (2016).

93. Fairfax, B.P. et al. Innate immune activity conditions the effect of regulatory variants upon monocyte gene expression. Science 343, 1246949 (2014).

94. Kerimov, N. et al. A compendium of uniformly processed human gene expression and splicing quantitative trait loci. Nat Genet 53, 1290–1299 (2021).

95. Mancuso, N. et al. Probabilistic fine-mapping of transcriptome-wide association studies. Nat Genet 51, 675–682 (2019).

96. Ioannidis, N.M. et al. REVEL: An Ensemble Method for Predicting the Pathogenicity of Rare Missense Variants. Am J Hum Genet 99, 877–885 (2016).

