## Supplementary Figures for "Deciphering Distinct Genetic Risk Factors for FTLD-TDP Pathological Subtypes via Whole-Genome Sequencing"

† In memoriam

*** Corresponding Authors**

Rosa Rademakers, Ph.D. (ORCID: 0000-0002-4049-0863)

VIB Center for Molecular Neurology

Universiteitsplein 1, 2610 Wilrijk, Belgium

Cyril Pottier, Ph.D.

Department of Neurology, Washington University School of Medicine

St Louis, MO, United States of America

[1]Department of Neuroscience, Mayo Clinic, 4500 San Pablo Road, Jacksonville, FL, 32224, United States of America

[2]Department of Biomedical Sciences, University of Antwerp, Antwerp, Belgium

[3]VIB Center for Molecular Neurology, VIB, Antwerp, Belgium

[4]Department of Neurology, Washington University School of Medicine, St Louis, MO, United States of America

[5]NeuroGenomics and Informatics Center, Washington University School of Medicine, St Louis, MO, United States of America

[6]Department of Quantitative Health Sciences, Mayo Clinic, Rochester, MN, 55905, United States of America

[7]Department of Neurology, Massachusetts General Hospital and Harvard Medical School, Boston, MA, 02114, United States of America

[8]Department of Neurology, Erasmus Medical Center, Wytemaweg 80, Rotterdam, 3015 CN, The Netherlands

[9]Department of Medicine, Division of Neurology, University of British Columbia, Vancouver, BC, V6T 2B5, Canada

[10]Division of Neurogeriatrics, Karolinska Institutet, Bioclinicum J10:20, Visionsgatan 4, Solna, 171 64, Sweden

[11]Unit for Hereditary Dementias, Karolinska University Hospital, Solna, 171 64, Sweden

[12]German Center for Neurodegenerative Diseases (DZNE), Tübingen, 72076, Germany

[13]Department of Neuropathology, University of Tübingen, Tübingen, 72076, Germany
[14]Department of Health Sciences Research, Mayo Clinic, 4500 San Pablo Road, Jacksonville, FL, 32224, United States of America
[15]Department of Neurology, Mayo Clinic, Rochester, MN, 55905, United States of America
[16]Department of Radiology, Mayo Clinic, Rochester, MN, 55905, United States of America
[17]University of Texas Health Science Center San Antonio, San Antonio, TX, 78229, United States of America
[18]Department of Neurology, UCSF Weill Institute for Neurosciences, University of California, San Francisco, San Francisco, CA, 94158, United States of America
[19]Department of Pathology, University of Utah, Salt Lake City, UT, 84112, United States of America
[20]Central Clinical School and Brain and Mind Centre, University of Sydney, Sydney, 2050, Australia
[21]University of Sydney, Sydney, 2050, Australia
[22]NeuRA, University of New South Wales, Randwick, 2031, Australia
[23]Department of Neurology, University of Washington, 325 9th Ave, Seattle, WA, 98104, United States of America
[24]Division Translational Genomics of Neurodegenerative Diseases, Center for Neurology and Hertie-Institute for Clinical Brain Research, University of Tübingen, Tübingen, Germany
[25]Department of Psychiatry and Behavioral Sciences, Johns Hopkins University, Baltimore, MD, 21218, United States of America
[26]Department of Neurology, Case Western Reserve University, Cleveland, OH, 44106, United States of America
[27]Division of Neuropathology, University of Texas Southwestern Medical Center, 5323 Harry Hines Blvd, Dallas, TX, 75390-9073, United States of America
[28]Department of Pathology and Laboratory Medicine, University of California, Davis Medical Center, Sacramento, CA, 95817, United States of America
[29]St. Michael's Hospital, 30 Bond St, Toronto, ON, M5B 1W8, Canada
[30]Department of Laboratory Medicine and Pathobiology, University of Toronto, Toronto, ON, M5S 1A1, Canada
[31]Sunnybrook Health Sciences Centre, Toronto, ON, M4N 3M5, Canada
[32]Krembil Discovery Tower, Tanz Centre for Research in Neurodegenerative Disease, University of Toronto, 60 Leonard Av, Toronto, ON, M5T 0S8, Canada
[33]Department of Pathology and Laboratory Medicine, Center for Neurodegenerative Disease Research,

Perelman School of Medicine at the University of Pennsylvania, Philadelphia, PA, 19104, United States of America
[34]Mesulam Center for Cognitive Neurology and Alzheimer's Disease, Northwestern University, Chicago, IL, 60611, United States of America
[35]Department of Psychiatry and Psychotherapy, Technical University of Munich, Munich, 80333, Germany
[36]kbo-Inn-Salzach-Klinikum, Clinical Center for Psychiatry, Psychotherapy, Psychosomatic Medicine, Geriatrics and Neurology, Wasserburg/Inn, 83512, Germany
[37]Department of Neurology, Indiana University School of Medicine, 355 West 16th Street, Indianapolis, IN, 46202, United States of America
[38]German Center for Neurodegenerative Diseases (DZNE), Feodor-Lynen-Str 17, Munich, 81377, Germany
[39]Department of Neurology, Mayo Clinic, Scottsdale, AZ, 85259, United States of America
[40]Department of Clinical Genetics, Erasmus Medical Center, Wytemaweg 80, Rotterdam, 3015 CN, The Netherlands
[41]Department of Psychiatry and Human Behavior, Brown Alpert Medical School, Brown University, Providence, RI, 02912, United States of America
[42]Banner Alzheimer's Institute, Phoenix, AZ, 85006, United States of America
[43]MRC Prion Unit at University College London, Institute of Prion Diseases, London, United Kingdom
[44]Department of Basic and Clinical Neuroscience, London Neurodegenerative Diseases Brain Bank, Institute of Psychiatry, Psychology and Neuroscience, King's College London, London, SE5 8AF, United Kingdom
[45]Department of Clinical Neuropathology, King's College Hospital NHS Foundation Trust, London, SE5 9RS, United Kingdom
[46]Centre for Neuropathology and Prion Research, Ludwig-Maximilians-University of Munich, Feodor-Lynen-Straße 23, Munich, 81377, Germany
[47]Department of Neurology, Mayo Clinic, 4500 San Pablo Road, Jacksonville, FL, 32224, United States of America
[48]Department of Neurology, David Geffen School of Medicine, University of California, Los Angeles, Los Angeles, CA, 90095, United States of America
[49]Department of Pathology, UCSF Weill Institute for Neurosciences, University of California, San Francisco, San Francisco, CA, 94158, United States of America

[50]School of Psychology and Brain and Mind Centre, University of Sydney, Sydney, 2050, Australia
[51]Department of Neurology, Penn Frontotemporal Degeneration Center, Perelman School of Medicine at the University of Pennsylvania, Philadelphia, PA, 19104, United States of America
[52]University of Washington BioRepository and Integrated Neuropathology (BRaIN) lab, Harborview Medical Center, 325 9th Ave, Seattle, WA, 98104, United States of America
[53]M.I.N.D. Institute Laboratory, University of California, Davis Medical Center, 2805 50th St, Sacramento, CA, 95817, United States of America
[54]German Center for Neurodegenerative Diseases (DZNE), Rostock, 18147, Germany
[55]Department of Neurology, Rostock University Medical Center, Rostock, 18147, Germany
[56]Alzheimer's Therapeutic Research Institute, Keck School of Medicine of the University of Southern California, San Diego, CA, USA
[57]Department of Psychiatry, Knight Alzheimer Disease Research Center, Washington University School of Medicine, Saint Louis, MO, 63108, United States of America
[58]Department of Pathology and Laboratory Medicine, Indiana University School of Medicine, 635 Barnhill Drive, Indianapolis, IN, 46202, United States of America
[59]Civin Laboratory for Neuropathology, Banner Sun Health Research Institute, Sun City, AZ, 85351, United States of America
[60]Department of Psychiatry and Psychotherapy, University Hospital, Ludwig-Maximilians-University of Munich, Munich, 81377, Germany
[61]Department of Neurology, Taub Institute for Research on Alzheimer's Disease and the Aging Brain, Columbia University Irving Medical Center, 630 West 168th St, New York, NY, 10032, United States of America
[62]Department of Pathology, Taub Institute for Research on Alzheimer's Disease and the Aging Brain, Columbia University Irving Medical Center, 630 West 168th St, New York, NY, 10032, United States of America
[63]Department of Neurology, University of Pittsburgh, Pittsburgh, PA, 15213, United States of America
[64]Department of Pathology, University of Pittsburgh, Pittsburgh, PA, 15213, United States of America
[65]Department of Pathology and Laboratory Medicine and Department of Neurology, Emory University, Atlanta, GA, 30322, United States of America
[66]Department of Neurodegenerative Disease, Dementia Research Centre, University College London Queen Square Institute of Neurology, London, WC1N 3BG, United Kingdom
[67]Department of Pathology, Feinberg School of Medicine, Northwestern University, Chicago, IL, 60611,

United States of America

[68]Department of Clinical Neurological Sciences, Schulich School of Medicine and Dentistry, University of Western Ontario, London, ON, N6A 2E2, Canada

[69]King's College Hospital NHS Foundation Trust, London, SE5 9RS, United Kingdom

[70]Department of Pathology and Laboratory Medicine, University of British Columbia, Vancouver, BC, V6T 1Z7, Canada

[71]Department of Psychiatry & Psychology, Mayo Clinic, Rochester, MN, 55905, United States of America

### SUPPLEMENTARY FIGURES

**Supplementary Figure 1. Age at onset and age at death distribution.** Violin plots showing age at onset (A) and age at death (B) are represented as well as box plots for each patient/control groups.

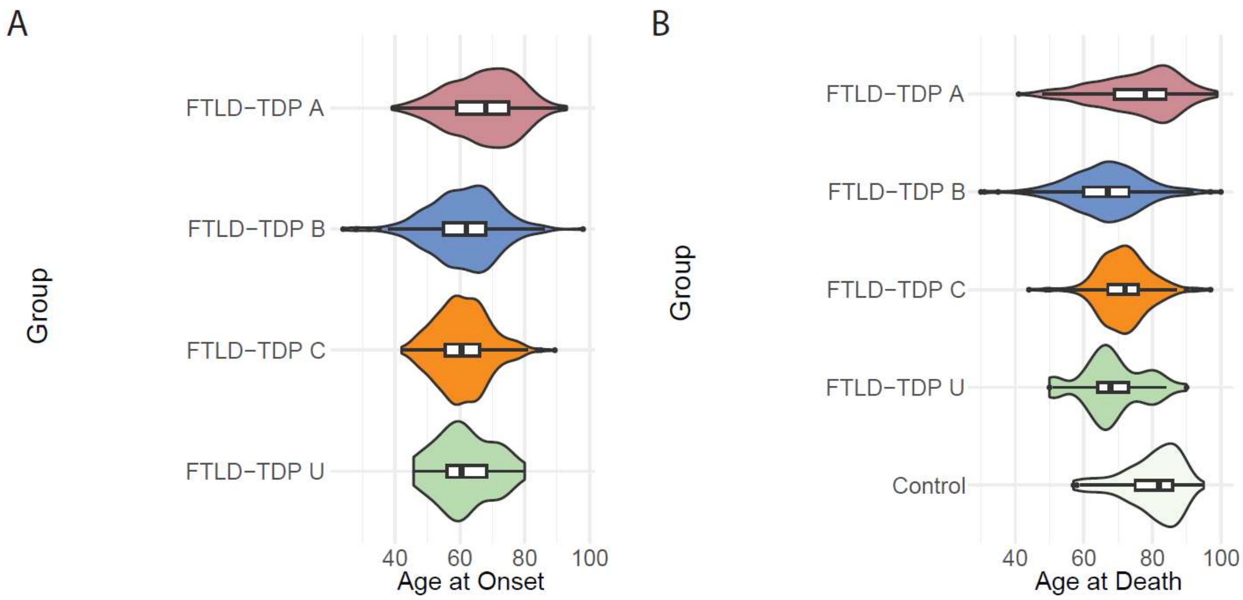

**Supplementary Figure 2. Gene prioritization results for FTLD-TDP All**. A visual summary of weighted evidence category scores for the prioritized genes within genome-wide significant and subthreshold loci for FTLD-TDP All GWAS. The leftmost squares indicate the locus index numbers which contain additional "_S" patterns for the subthreshold loci, meanwhile others indicate the genome-wide significant loci. The types of evidence for each category are colored according to the six different domains to which they belonged. Weighted scores for each evidence category are rescaled to a 0–100 scale based on the maximum score a candidate gene can obtain from a category (see **Supplementary Table 2**). The darker colors represent higher scores in categories, while tier 1 prioritized genes are displayed in dark green and tier 2 prioritized genes are displayed in light green. Only tier 1 and tier 2 genes are shown for each locus, and all candidate genes considered and scored can be found in **Supplementary Table 3**. MAFs (based on gnomAD v4 non-Finnish European samples) and CADD (v1.7) PHRED scores for rare and/or protein-altering rare variants are labeled in white within the respective squares. eQTL, expression QTL; sQTL, splicing QTL; mQTL, methylation QTL; pQTL, protein-expression QTL; haQTL, histone acetylation QTL; coloc, colocalization; eTWAS, expression transcriptome-wide association study; sTWAS, splicing transcriptome-wide association study; PWAS, proteome-wide association study; Mon. Mac., monocytes and macrophages; LCL, lymphoblastoid cell line; QTLCat, The eQTL Catalogue.

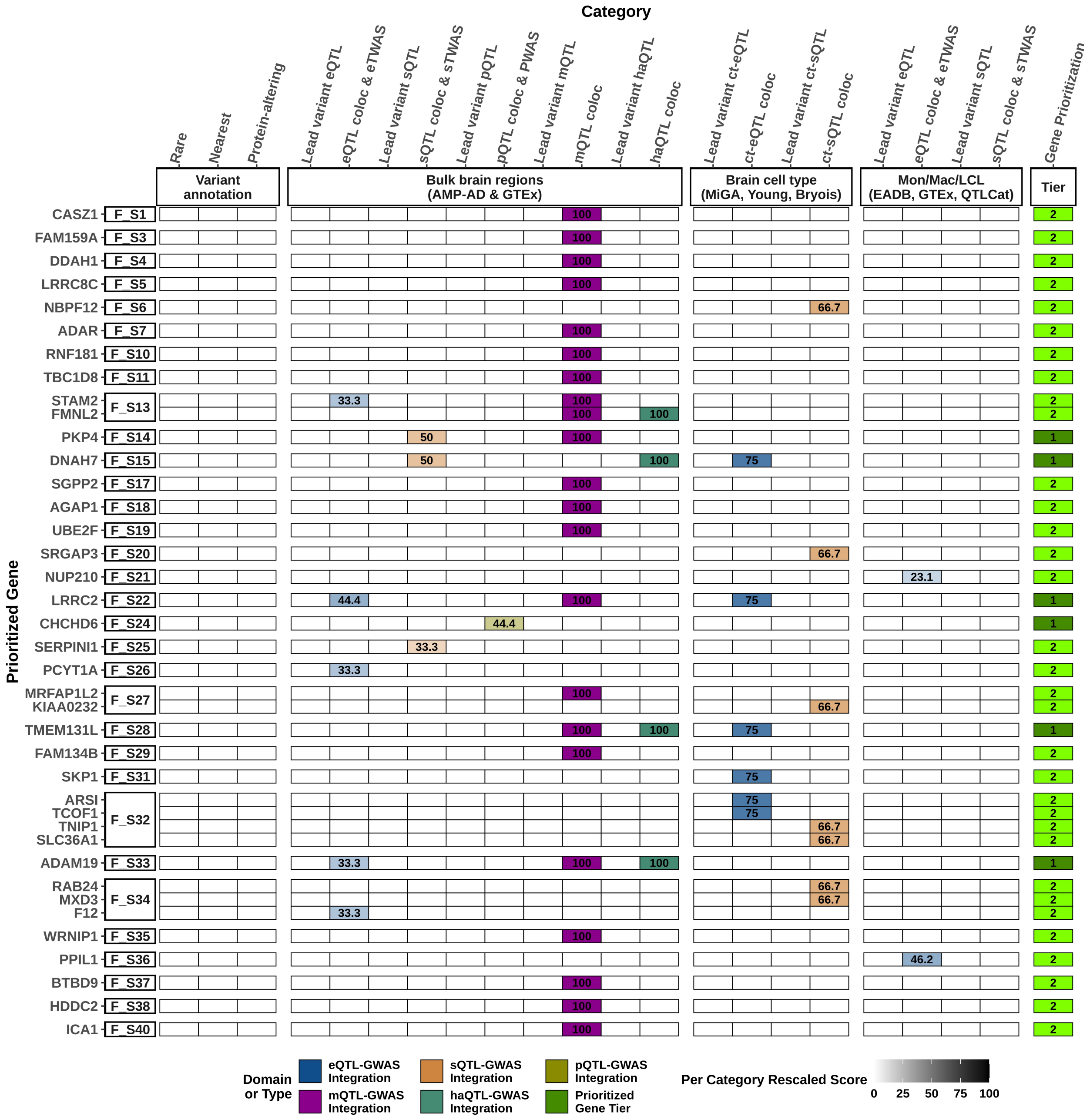

Category
Rare
Nearest
Protein-altering
Lead variant eQTL
eQTL coloc & eTWAS
Lead variant sQTL
sQTL coloc & sTWAS
Lead variant pQTL
pQTL coloc & PWAS
Lead variant mQTL
mQTL coloc
Lead variant haQTL
haQTL coloc
Lead variant ct-eQTL
ct-eQTL coloc
Lead variant ct-sQTL
ct-sQTL coloc
Lead variant eQTL
eQTL coloc & eTWAS
Lead variant sQTL
sQTL coloc & sTWAS
Gene Prioritization
Variant annotation
Bulk brain regions (AMP-AD & GTEx)
Brain cell type (MiGA, Young, Bryois)
Mon/Mac/LCL (EADB, GTEx, QTLCat)
Tier
Prioritized Gene
CASZ1 F_S1
FAM159A F_S3
DDAH1 F_S4
LRRC8C F_S5
NBPF12 F_S6
ADAR F_S7
RNF181 F_S10
TBC1D8 F_S11
STAM2 FMNL2 F_S13
PKP4 F_S14
DNAH7 F_S15
SGPP2 F_S17
AGAP1 F_S18
UBE2F F_S19
SRGAP3 F_S20
NUP210 F_S21
LRRC2 F_S22
CHCHD6 F_S24
SERPINI1 F_S25
PCYT1A F_S26
MRFAP1L2 KIAA0232 F_S27
TMEM131L F_S28
FAM134B F_S29
SKP1 F_S31
ARSI TCOF1 TNIP1 SLC36A1 F_S32
ADAM19 F_S33
RAB24 MXD3 F12 F_S34
WRNIP1 F_S35
PPIL1 F_S36
BTBD9 F_S37
HDDC2 F_S38
ICA1 F_S40
Domain or Type
eQTL-GWAS Integration
mQTL-GWAS Integration
sQTL-GWAS Integration
haQTL-GWAS Integration
pQTL-GWAS Integration
Prioritized Gene Tier
Per Category Rescaled Score
0 25 50 75 100

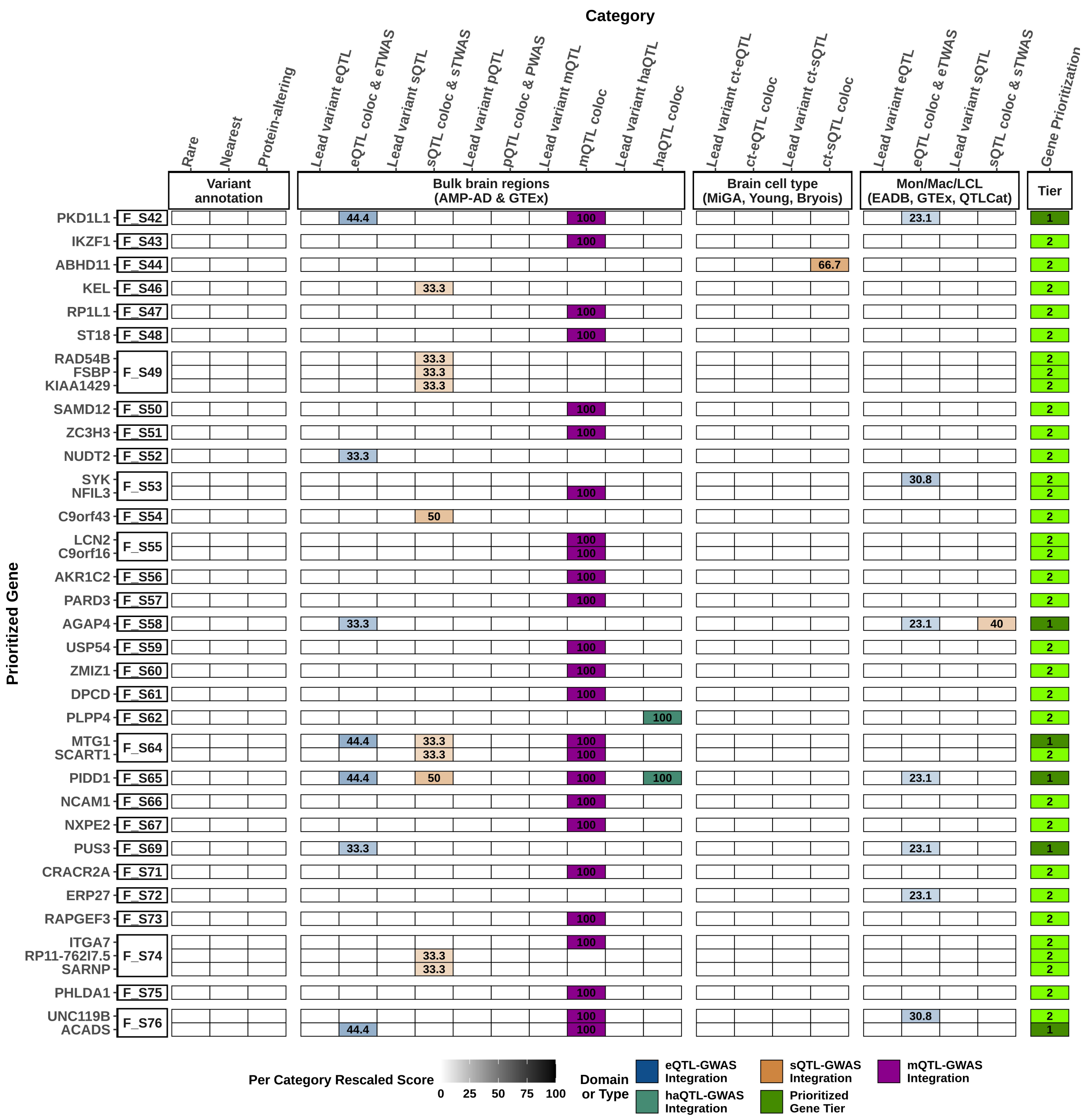

Category
Rare
Nearest
Protein-altering
Lead variant eQTL
eQTL coloc & eTWAS
Lead variant sQTL
sQTL coloc & sTWAS
Lead variant pQTL
pQTL coloc & PWAS
Lead variant mQTL
mQTL coloc
Lead variant haQTL
haQTL coloc
Lead variant ct-eQTL
ct-eQTL coloc
Lead variant ct-sQTL
ct-sQTL coloc
Lead variant eQTL
eQTL coloc & eTWAS
Lead variant sQTL
sQTL coloc & sTWAS
Gene Prioritization
Variant annotation
Bulk brain regions (AMP-AD & GTEx)
Brain cell type (MiGA, Young, Bryois)
Mon/Mac/LCL (EADB, GTEx, QTLCat)
Tier
Prioritized Gene
PKD1L1 F_S42 44.4 100 23.1 1
IKZF1 F_S43 100 2
ABHD11 F_S44 66.7 2
KEL F_S46 33.3 2
RP1L1 F_S47 100 2
ST18 F_S48 100 2
RAD54B F_S49 33.3 2
FSBP 33.3 2
KIAA1429 33.3 2
SAMD12 F_S50 100 2
ZC3H3 F_S51 100 2
NUDT2 F_S52 33.3 2
SYK F_S53 30.8 2
NFIL3 100 2
C9orf43 F_S54 50 2
LCN2 F_S55 100 2
C9orf16 100 2
AKR1C2 F_S56 100 2
PARD3 F_S57 100 2
AGAP4 F_S58 33.3 23.1 40 1
USP54 F_S59 100 2
ZMIZ1 F_S60 100 2
DPCD F_S61 100 2
PLPP4 F_S62 100 2
MTG1 F_S64 44.4 33.3 100 1
SCART1 33.3 100 2
PIDD1 F_S65 44.4 50 100 100 23.1 1
NCAM1 F_S66 100 2
NXPE2 F_S67 100 2
PUS3 F_S69 33.3 23.1 1
CRACR2A F_S71 100 2
ERP27 F_S72 23.1 2
RAPGEF3 F_S73 100 2
ITGA7 F_S74 100 2
RP11-762I7.5 33.3 2
SARNP 33.3 2
PHLDA1 F_S75 100 2
UNC119B F_S76 100 30.8 2
ACADS 44.4 100 1
Per Category Rescaled Score
0 25 50 75 100
Domain or Type
eQTL-GWAS Integration
sQTL-GWAS Integration
mQTL-GWAS Integration
haQTL-GWAS Integration
Prioritized Gene Tier

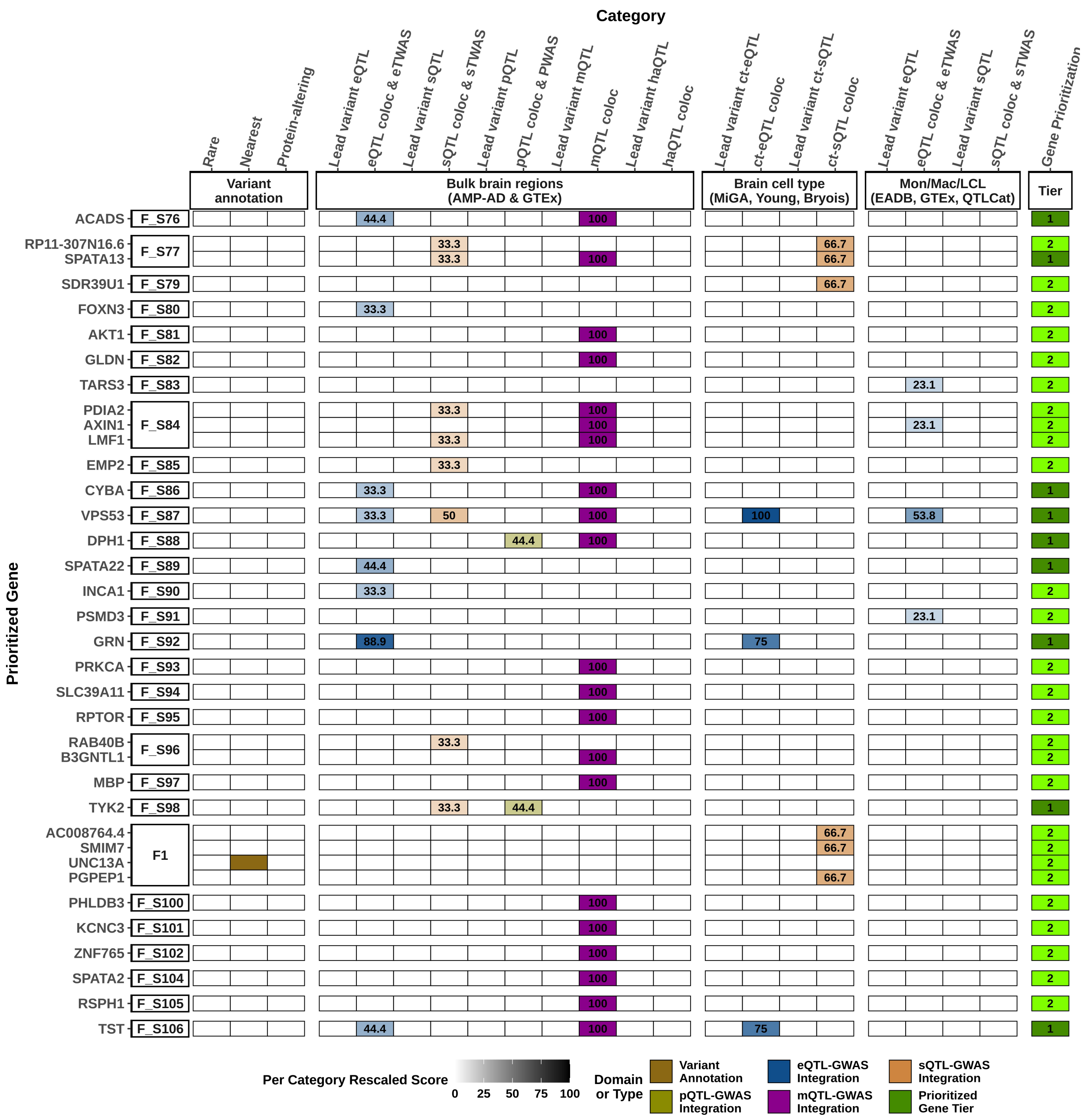

Category
Rare
Nearest
Protein-altering
Lead variant eQTL
eQTL coloc & eTWAS
Lead variant sQTL
sQTL coloc & sTWAS
Lead variant pQTL
pQTL coloc & PWAS
Lead variant mQTL
mQTL coloc
Lead variant haQTL
haQTL coloc
Lead variant ct-eQTL
ct-eQTL coloc
Lead variant ct-sQTL
ct-sQTL coloc
Lead variant eQTL
eQTL coloc & eTWAS
Lead variant sQTL
sQTL coloc & sTWAS
Gene Prioritization
Variant annotation
Bulk brain regions (AMP-AD & GTEx)
Brain cell type (MiGA, Young, Bryois)
Mon/Mac/LCL (EADB, GTEx, QTLCat)
Tier
Prioritized Gene
ACADS F_S76
RP11-307N16.6
SPATA13 F_S77
SDR39U1 F_S79
FOXN3 F_S80
AKT1 F_S81
GLDN F_S82
TARS3 F_S83
PDIA2
AXIN1
LMF1 F_S84
EMP2 F_S85
CYBA F_S86
VPS53 F_S87
DPH1 F_S88
SPATA22 F_S89
INCA1 F_S90
PSMD3 F_S91
GRN F_S92
PRKCA F_S93
SLC39A11 F_S94
RPTOR F_S95
RAB40B
B3GNTL1 F_S96
MBP F_S97
TYK2 F_S98
AC008764.4
SMIM7
UNC13A
PGPEP1 F1
PHLDB3 F_S100
KCNC3 F_S101
ZNF765 F_S102
SPATA2 F_S104
RSPH1 F_S105
TST F_S106
Per Category Rescaled Score
0 25 50 75 100
Domain or Type
Variant Annotation
eQTL-GWAS Integration
sQTL-GWAS Integration
pQTL-GWAS Integration
mQTL-GWAS Integration
Prioritized Gene Tier

**Supplementary Figure 3. Gene prioritization results for FTLD-TDP A**. A visual summary of weighted evidence category scores for the prioritized genes within genome-wide significant and subthreshold loci for FTLD-TDP A GWAS. The leftmost squares indicate the locus index numbers which contain additional "_S" patterns for the subthreshold loci, meanwhile others indicate the genome-wide significant loci. The types of evidence for each category are colored according to the six different domains to which they belonged. Weighted scores for each evidence category are rescaled to a 0–100 scale based on the maximum score a candidate gene can obtain from a category (see **Supplementary Table 2**). The darker colors represent higher scores in categories, while tier 1 prioritized genes are displayed in dark green and tier 2 prioritized genes are displayed in light green. Only tier 1 and tier 2 genes are shown for each locus, and all candidate genes considered and scored can be found in **Supplementary Table 3**. MAFs (based on gnomAD v4 non-Finnish European samples) and CADD (v1.7) PHRED scores for rare and/or protein-altering rare variants are labeled in white within the respective squares. eQTL, expression QTL; sQTL, splicing QTL; mQTL, methylation QTL; pQTL, protein-expression QTL; haQTL, histone acetylation QTL; coloc, colocalization; eTWAS, expression transcriptome-wide association study; sTWAS, splicing transcriptome-wide association study; PWAS, proteome-wide association study; Mon. Mac., monocytes and macrophages; LCL, lymphoblastoid cell line; QTLCat, The eQTL Catalogue.

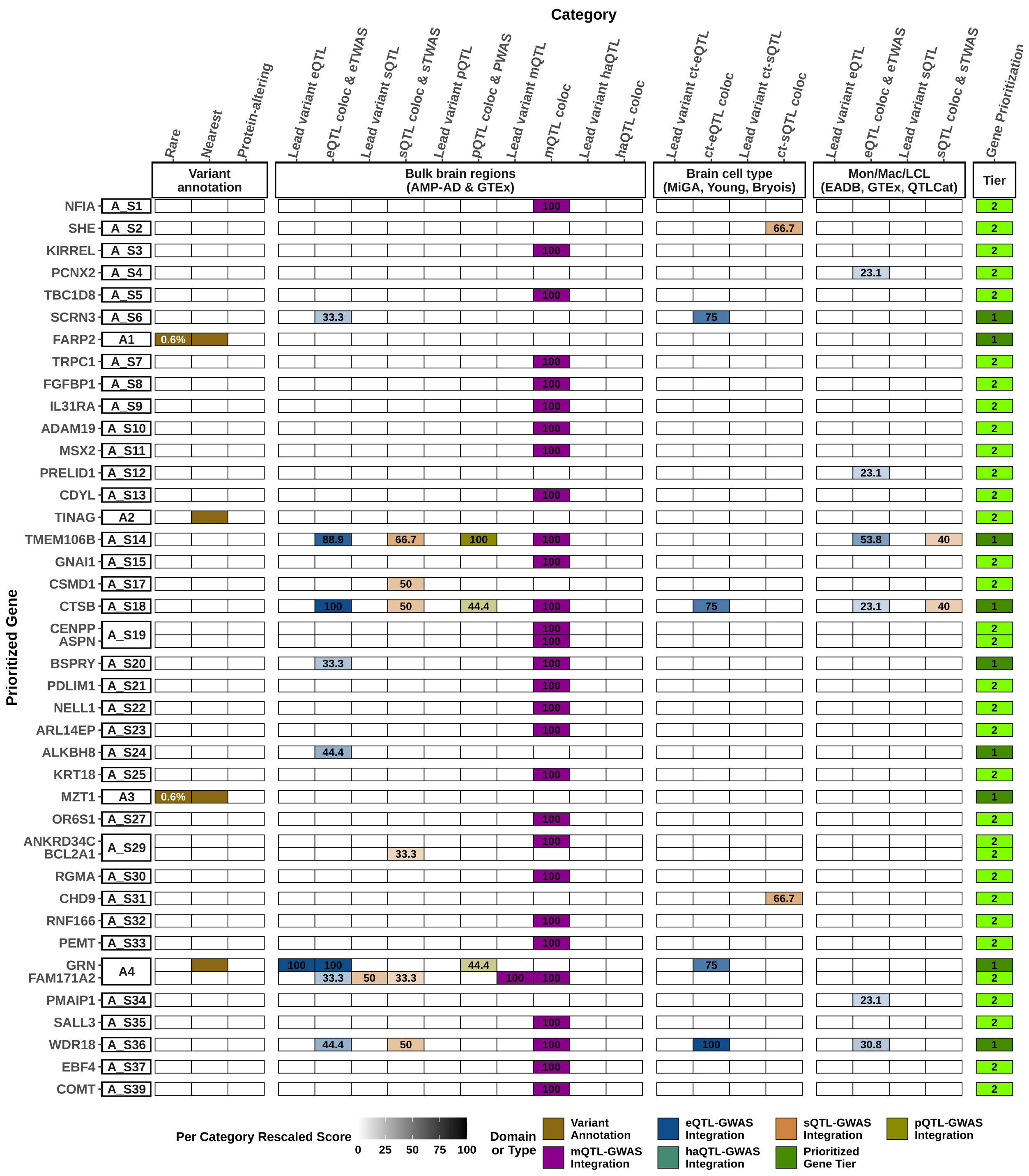

Category
Rare
Nearest
Protein-altering
Lead variant eQTL
eQTL coloc & eTWAS
Lead variant sQTL
sQTL coloc & sTWAS
Lead variant pQTL
pQTL coloc & PWAS
Lead variant mQTL
mQTL coloc
Lead variant haQTL
haQTL coloc
Lead variant ct-eQTL
ct-eQTL coloc
Lead variant ct-sQTL
ct-sQTL coloc
Lead variant eQTL
eQTL coloc & eTWAS
Lead variant sQTL
sQTL coloc & sTWAS
Gene Prioritization
Variant annotation
Bulk brain regions (AMP-AD & GTEx)
Brain cell type (MiGA, Young, Bryois)
Mon/Mac/LCL (EADB, GTEx, QTLCat)
Tier
Prioritized Gene
NFIA A_S1
SHE A_S2
KIRREL A_S3
PCNX2 A_S4
TBC1D8 A_S5
SCRN3 A_S6
FARP2 A1
TRPC1 A_S7
FGFBP1 A_S8
IL31RA A_S9
ADAM19 A_S10
MSX2 A_S11
PRELID1 A_S12
CDYL A_S13
TINAG A2
TMEM106B A_S14
GNAI1 A_S15
CSMD1 A_S17
CTSB A_S18
CENPP ASPN A_S19
BSPRY A_S20
PDLIM1 A_S21
NELL1 A_S22
ARL14EP A_S23
ALKBH8 A_S24
KRT18 A_S25
MZT1 A3
OR6S1 A_S27
ANKRD34C BCL2A1 A_S29
RGMA A_S30
CHD9 A_S31
RNF166 A_S32
PEMT A_S33
GRN FAM171A2 A4
PMAIP1 A_S34
SALL3 A_S35
WDR18 A_S36
EBF4 A_S37
COMT A_S39
Per Category Rescaled Score
0 25 50 75 100
Domain or Type
Variant Annotation
eQTL-GWAS Integration
sQTL-GWAS Integration
pQTL-GWAS Integration
mQTL-GWAS Integration
haQTL-GWAS Integration
Prioritized Gene Tier

**Supplementary Figure 4. Gene prioritization results for FTLD-TDP B**. A visual summary of weighted evidence category scores for the prioritized genes within genome-wide significant and subthreshold loci for FTLD-TDP B GWAS. The leftmost squares indicate the locus index numbers which contain additional "_S" patterns for the subthreshold loci, meanwhile others indicate the genome-wide significant loci. The types of evidence for each category are colored according to the six different domains to which they belonged. Weighted scores for each evidence category are rescaled to a 0–100 scale based on the maximum score a candidate gene can obtain from a category (see **Supplementary Table 2**). The darker colors represent higher scores in categories, while tier 1 prioritized genes are displayed in dark green and tier 2 prioritized genes are displayed in light green. Only tier 1 and tier 2 genes are shown for each locus, and all candidate genes considered and scored can be found in **Supplementary Table 3**. MAFs (based on gnomAD v4 non-Finnish European samples) and CADD (v1.7) PHRED scores for rare and/or protein-altering rare variants are labeled in white within the respective squares. eQTL, expression QTL; sQTL, splicing QTL; mQTL, methylation QTL; pQTL, protein-expression QTL; haQTL, histone acetylation QTL; coloc, colocalization; eTWAS, expression transcriptome-wide association study; sTWAS, splicing transcriptome-wide association study; PWAS, proteome-wide association study; Mon. Mac., monocytes and macrophages; LCL, lymphoblastoid cell line; QTLCat, The eQTL Catalogue.

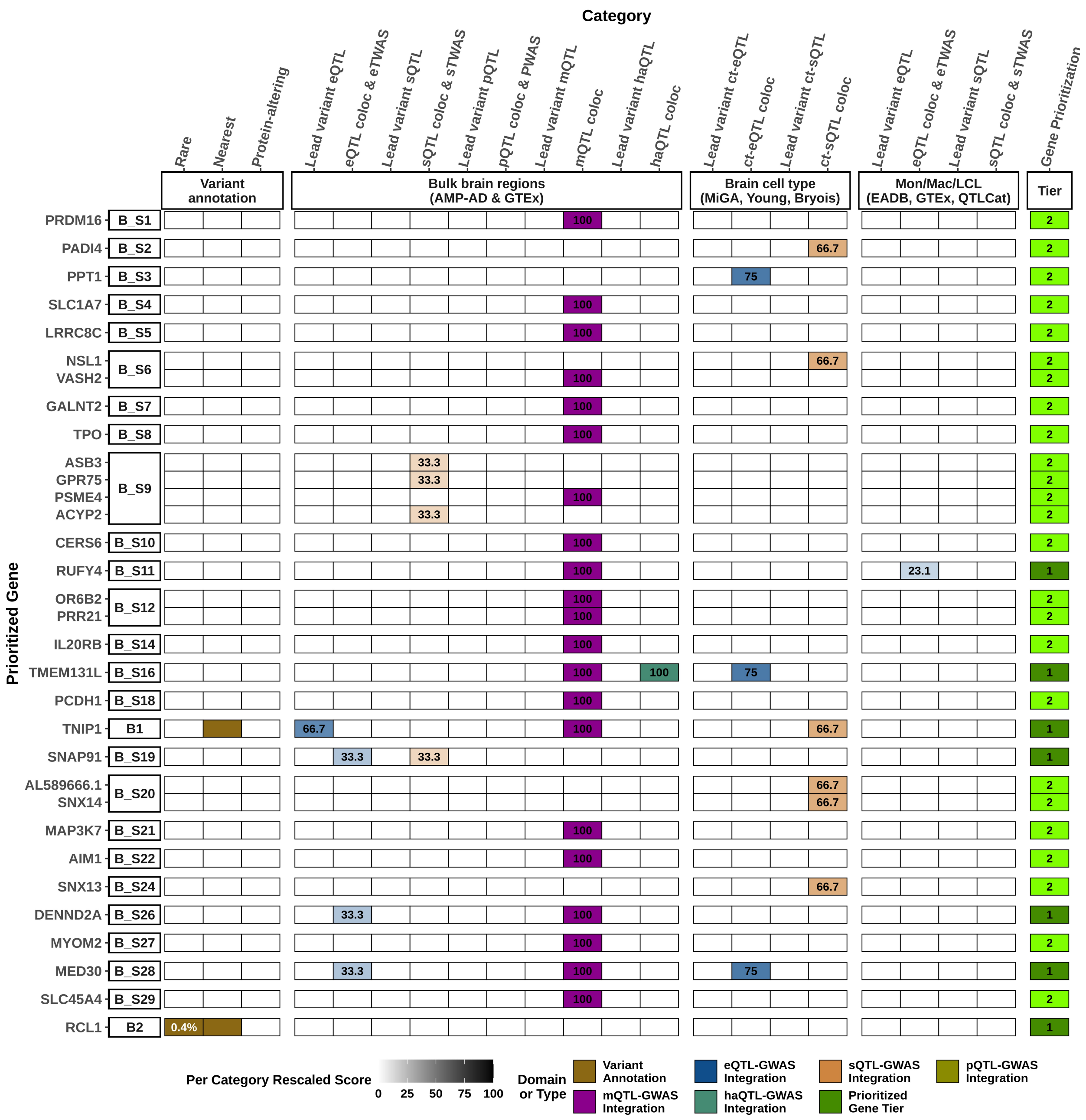

Category
Rare
Nearest
Protein-altering
Lead variant eQTL
eQTL coloc & eTWAS
Lead variant sQTL
sQTL coloc & sTWAS
Lead variant pQTL
pQTL coloc & PWAS
Lead variant mQTL
mQTL coloc
Lead variant haQTL
haQTL coloc
Lead variant ct-eQTL
ct-eQTL coloc
Lead variant ct-sQTL
ct-sQTL coloc
Lead variant eQTL
eQTL coloc & eTWAS
Lead variant sQTL
sQTL coloc & sTWAS
Gene Prioritization
Variant annotation
Bulk brain regions (AMP-AD & GTEx)
Brain cell type (MiGA, Young, Bryois)
Mon/Mac/LCL (EADB, GTEx, QTLCat)
Tier
Prioritized Gene
PRDM16 B_S1
PADI4 B_S2
PPT1 B_S3
SLC1A7 B_S4
LRRC8C B_S5
NSL1
VASH2 B_S6
GALNT2 B_S7
TPO B_S8
ASB3
GPR75
PSME4
ACYP2 B_S9
CERS6 B_S10
RUFY4 B_S11
OR6B2
PRR21 B_S12
IL20RB B_S14
TMEM131L B_S16
PCDH1 B_S18
TNIP1 B1
SNAP91 B_S19
AL589666.1
SNX14 B_S20
MAP3K7 B_S21
AIM1 B_S22
SNX13 B_S24
DENND2A B_S26
MYOM2 B_S27
MED30 B_S28
SLC45A4 B_S29
RCL1 B2
100
66.7
75
33.3
23.1
0.4%
1
2
Per Category Rescaled Score
0 25 50 75 100
Domain or Type
Variant Annotation
mQTL-GWAS Integration
eQTL-GWAS Integration
haQTL-GWAS Integration
sQTL-GWAS Integration
Prioritized Gene Tier
pQTL-GWAS Integration

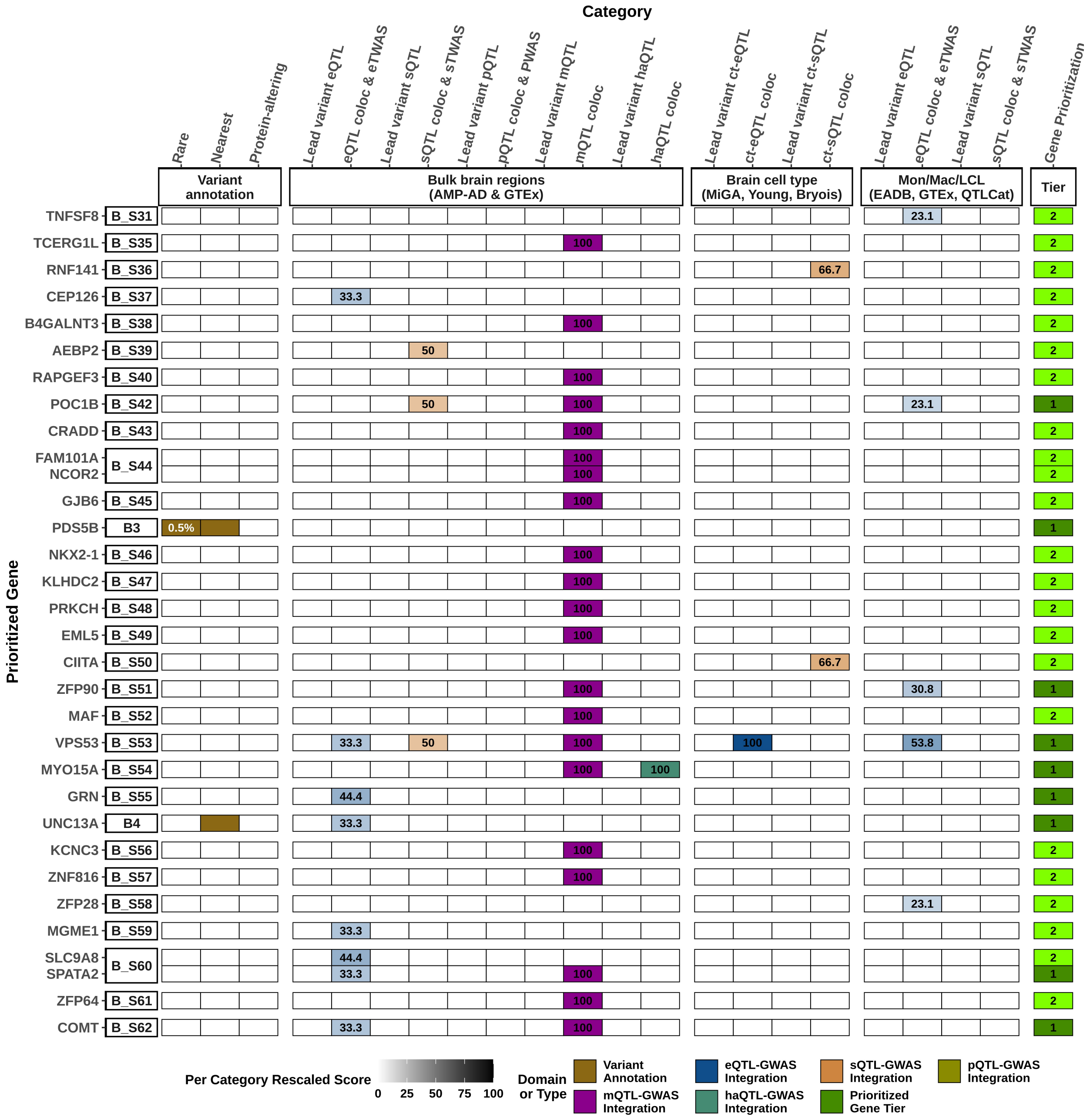
Category
Rare
Nearest
Protein-altering
Lead variant eQTL
eQTL coloc & eTWAS
Lead variant sQTL
sQTL coloc & sTWAS
Lead variant pQTL
pQTL coloc & PWAS
Lead variant mQTL
mQTL coloc
Lead variant haQTL
haQTL coloc
Lead variant ct-eQTL
ct-eQTL coloc
Lead variant ct-sQTL
ct-sQTL coloc
Lead variant eQTL
eQTL coloc & eTWAS
Lead variant sQTL
sQTL coloc & sTWAS
Gene Prioritization
Variant annotation
Bulk brain regions (AMP-AD & GTEx)
Brain cell type (MiGA, Young, Bryois)
Mon/Mac/LCL (EADB, GTEx, QTLCat)
Tier
Prioritized Gene
TNFSF8 B_S31 23.1 2
TCERG1L B_S35 100 2
RNF141 B_S36 66.7 2
CEP126 B_S37 33.3 2
B4GALNT3 B_S38 100 2
AEBP2 B_S39 50 2
RAPGEF3 B_S40 100 2
POC1B B_S42 50 100 23.1 1
CRADD B_S43 100 2
FAM101A B_S44 100 2
NCOR2 100 2
GJB6 B_S45 100 2
PDS5B B3 0.5% 1
NKX2-1 B_S46 100 2
KLHDC2 B_S47 100 2
PRKCH B_S48 100 2
EML5 B_S49 100 2
CIITA B_S50 66.7 2
ZFP90 B_S51 100 30.8 1
MAF B_S52 100 2
VPS53 B_S53 33.3 50 100 100 53.8 1
MYO15A B_S54 100 100 1
GRN B_S55 44.4 1
UNC13A B4 33.3 1
KCNC3 B_S56 100 2
ZNF816 B_S57 100 2
ZFP28 B_S58 23.1 2
MGME1 B_S59 33.3 2
SLC9A8 B_S60 44.4 2
SPATA2 33.3 100 1
ZFP64 B_S61 100 2
COMT B_S62 33.3 100 1
Per Category Rescaled Score
0 25 50 75 100
Domain or Type
Variant Annotation
mQTL-GWAS Integration
eQTL-GWAS Integration
haQTL-GWAS Integration
sQTL-GWAS Integration
Prioritized Gene Tier
pQTL-GWAS Integration

**Supplementary Figure 5. Gene prioritization results for FTLD-TDP C**. A visual summary of weighted evidence category scores for the prioritized genes within genome-wide significant and subthreshold loci for FTLD-TDP C GWAS. The leftmost squares indicate the locus index numbers which contain additional "_S" patterns for the subthreshold loci, meanwhile others indicate the genome-wide significant loci. The types of evidence for each category are colored according to the six different domains to which they belonged. Weighted scores for each evidence category are rescaled to a 0–100 scale based on the maximum score a candidate gene can obtain from a category (see **Supplementary Table 2**). The darker colors represent higher scores in categories, while tier 1 prioritized genes are displayed in dark green and tier 2 prioritized genes are displayed in light green. Only tier 1 and tier 2 genes are shown for each locus, and all candidate genes considered and scored can be found in **Supplementary Table 3**. MAFs (based on gnomAD v4 non-Finnish European samples) and CADD (v1.7) PHRED scores for rare and/or protein-altering rare variants are labeled in white within the respective squares. eQTL, expression QTL; sQTL, splicing QTL; mQTL, methylation QTL; pQTL, protein-expression QTL; haQTL, histone acetylation QTL; coloc, colocalization; eTWAS, expression transcriptome-wide association study; sTWAS, splicing transcriptome-wide association study; PWAS, proteome-wide association study; Mon. Mac., monocytes and macrophages; LCL, lymphoblastoid cell line; QTLCat, The eQTL Catalogue.

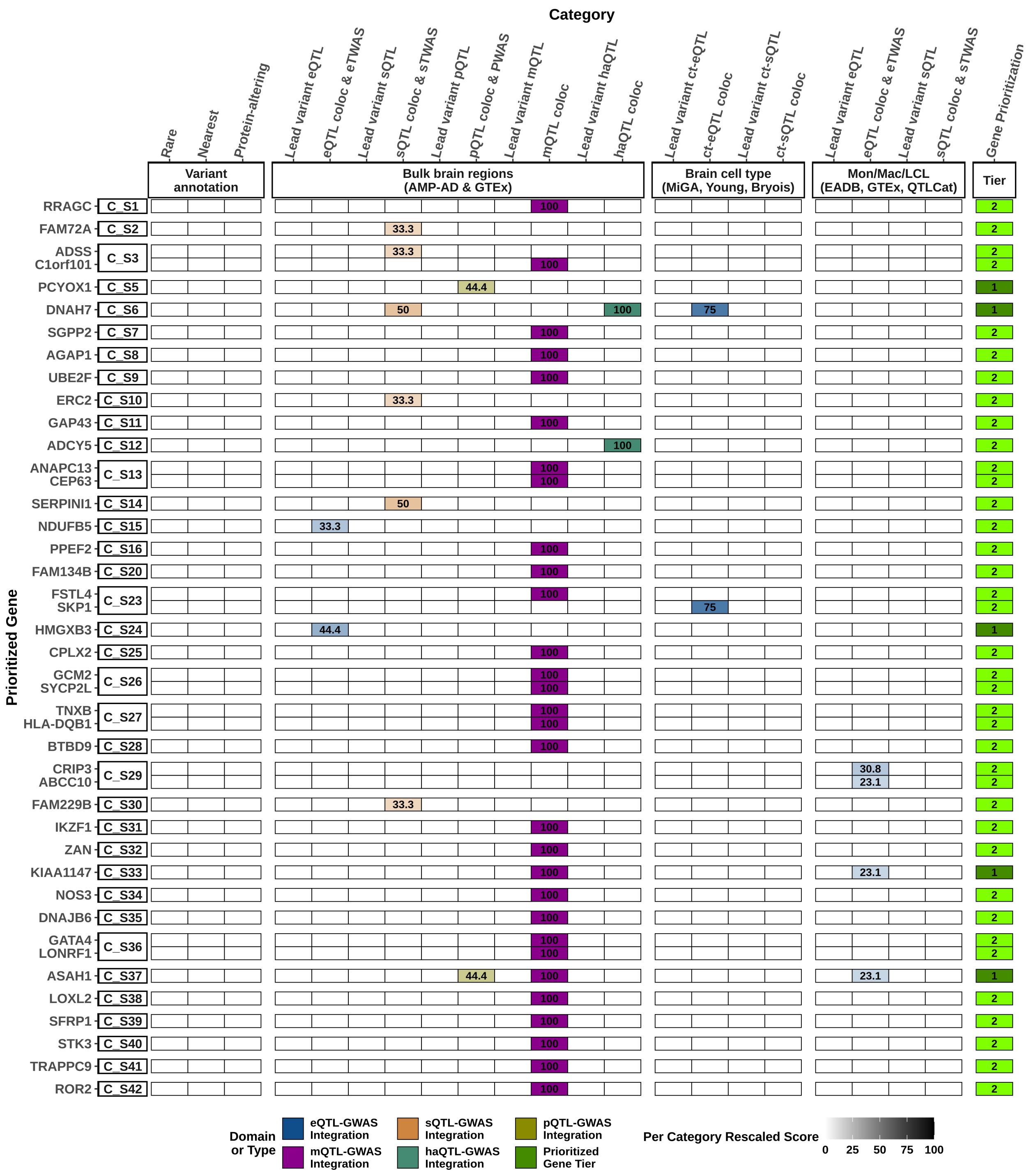
Category
Rare
Nearest
Protein-altering
Lead variant eQTL
eQTL coloc & eTWAS
Lead variant sQTL
sQTL coloc & sTWAS
Lead variant pQTL
pQTL coloc & PWAS
Lead variant mQTL
mQTL coloc
Lead variant haQTL
haQTL coloc
Lead variant ct-eQTL
ct-eQTL coloc
Lead variant ct-sQTL
ct-sQTL coloc
Lead variant eQTL
eQTL coloc & eTWAS
Lead variant sQTL
sQTL coloc & sTWAS
Gene Prioritization
Variant annotation
Bulk brain regions (AMP-AD & GTEx)
Brain cell type (MiGA, Young, Bryois)
Mon/Mac/LCL (EADB, GTEx, QTLCat)
Tier
Prioritized Gene
RRAGC C_S1
FAM72A C_S2
ADSS C1orf101 C_S3
PCYOX1 C_S5
DNAH7 C_S6
SGPP2 C_S7
AGAP1 C_S8
UBE2F C_S9
ERC2 C_S10
GAP43 C_S11
ADCY5 C_S12
ANAPC13 CEP63 C_S13
SERPINI1 C_S14
NDUFB5 C_S15
PPEF2 C_S16
FAM134B C_S20
FSTL4 SKP1 C_S23
HMGXB3 C_S24
CPLX2 C_S25
GCM2 SYCP2L C_S26
TNXB HLA-DQB1 C_S27
BTBD9 C_S28
CRIP3 ABCC10 C_S29
FAM229B C_S30
IKZF1 C_S31
ZAN C_S32
KIAA1147 C_S33
NOS3 C_S34
DNAJB6 C_S35
GATA4 LONRF1 C_S36
ASAH1 C_S37
LOXL2 C_S38
SFRP1 C_S39
STK3 C_S40
TRAPPC9 C_S41
ROR2 C_S42
Domain or Type
eQTL-GWAS Integration
sQTL-GWAS Integration
pQTL-GWAS Integration
mQTL-GWAS Integration
haQTL-GWAS Integration
Prioritized Gene Tier
Per Category Rescaled Score
0 25 50 75 100

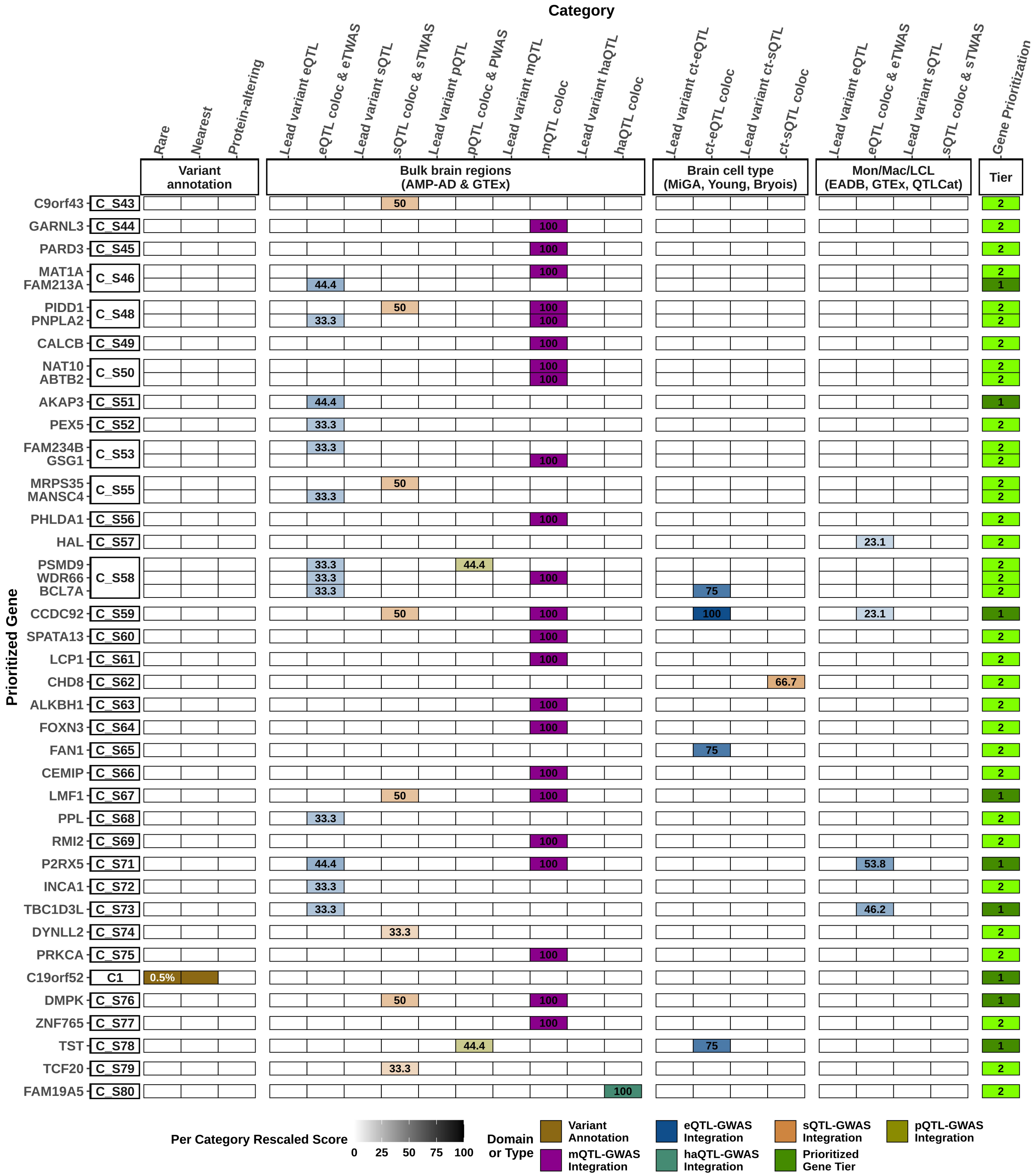
Category
Rare
Nearest
Protein-altering
Lead variant eQTL
eQTL coloc & eTWAS
Lead variant sQTL
sQTL coloc & sTWAS
Lead variant pQTL
pQTL coloc & PWAS
Lead variant mQTL
mQTL coloc
Lead variant haQTL
haQTL coloc
Lead variant ct-eQTL
ct-eQTL coloc
Lead variant ct-sQTL
ct-sQTL coloc
Lead variant eQTL
eQTL coloc & eTWAS
Lead variant sQTL
sQTL coloc & sTWAS
Gene Prioritization
Variant annotation
Bulk brain regions (AMP-AD & GTEx)
Brain cell type (MiGA, Young, Bryois)
Mon/Mac/LCL (EADB, GTEx, QTLCat)
Tier
Prioritized Gene
C9orf43 C_S43
GARNL3 C_S44
PARD3 C_S45
MAT1A FAM213A C_S46
PIDD1 PNPLA2 C_S48
CALCB C_S49
NAT10 ABTB2 C_S50
AKAP3 C_S51
PEX5 C_S52
FAM234B GSG1 C_S53
MRPS35 MANSC4 C_S55
PHLDA1 C_S56
HAL C_S57
PSMD9 WDR66 BCL7A C_S58
CCDC92 C_S59
SPATA13 C_S60
LCP1 C_S61
CHD8 C_S62
ALKBH1 C_S63
FOXN3 C_S64
FAN1 C_S65
CEMIP C_S66
LMF1 C_S67
PPL C_S68
RMI2 C_S69
P2RX5 C_S71
INCA1 C_S72
TBC1D3L C_S73
DYNLL2 C_S74
PRKCA C_S75
C19orf52 C1
DMPK C_S76
ZNF765 C_S77
TST C_S78
TCF20 C_S79
FAM19A5 C_S80
0.5%
Per Category Rescaled Score
0 25 50 75 100
Domain or Type
Variant Annotation
eQTL-GWAS Integration
sQTL-GWAS Integration
pQTL-GWAS Integration
mQTL-GWAS Integration
haQTL-GWAS Integration
Prioritized Gene Tier

**Supplementary Figure 6. *RBPJL* and *L3MBTL1* RNA expression in brain tissue**. **A** Weighted gene co-expression network from the ROSMAP dataset and the BrainExp database showing that both genes are part of the same module. **B** Single nuclei RNA expression of RBPJL and L3MBTL1 genes from the Seattle Alzheimer's Disease Brain cell Atlas from middle temporal gyrus of 84 aged donors. The darkness of the color of each nucleus increases as the expression level becomes higher.

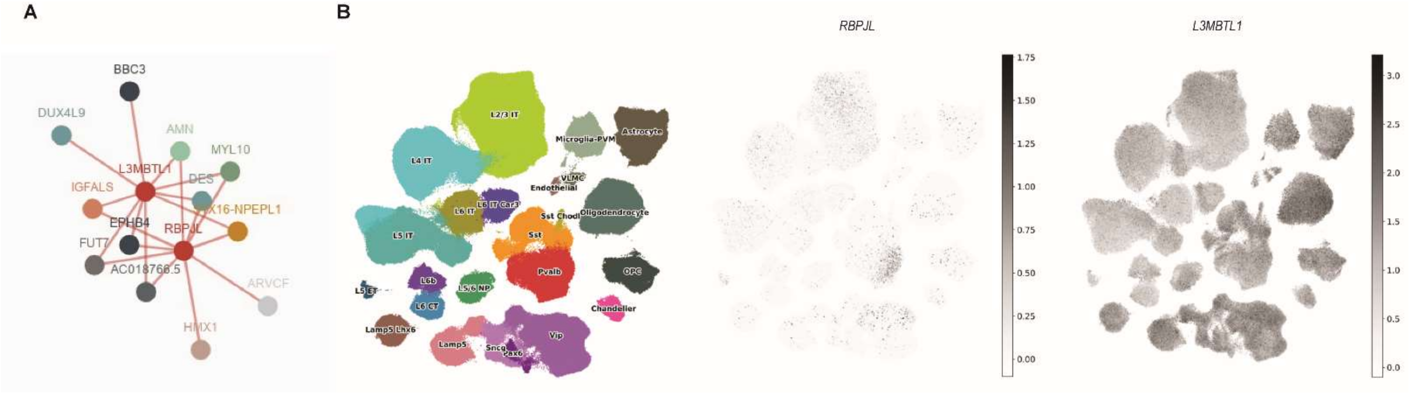
